# COVID-19 vaccine uptake in United States counties: geospatial vaccination patterns and trajectories towards herd immunity

**DOI:** 10.1101/2021.05.28.21257946

**Authors:** Pavel Chernyavskiy, Jeanita W. Richardson, Sarah J. Ratcliffe

## Abstract

Following the COVID-19 pandemic, safe and effective vaccines were developed and authorized for use in the general population. Studying factors that encourage community acceptance of these vaccines is needed to prevent proliferation of SARS-CoV-2 variants, to safely relax local restrictions, and to return to pre-pandemic living conditions. To our knowledge, United States (US) county-level disparities in vaccination are yet to be investigated. Our data span February - May 2021 across 3138 US counties. We consider percentage of residents with at least one dose of an authorized COVID vaccine as the outcome. Spatio-temporal models were used to determine associations of vaccination rates with time-fixed and time-varying covariates. Spatial variability was modelled via Conditional Auto-regressive models; county trajectories over time were specified using random slopes. Greater vaccination rates occur in counties with older residents, high educational attainment, and high proportion of minority residents. Vaccination rates change with COVID risk metrics, suggesting continued slowing of vaccine uptake due to decreasing incidence and infection rates. County effects reveal strong regional patterns in average vaccination rates and trajectories. Although local herd immunity can be expected in August 2021 for counties with typical uptake rates, these counties are clustered in relatively few areas of the country.

## INTRODUCTION

The Coronavirus Disease (COVID)-19 pandemic has been a defining generational event, with no country, region, or municipality spared. Following the rapid development of several safe and effective vaccines^1-3^, encouraging broad community acceptance has become crucially important^4,5^. Although some progress towards population herd immunity is expected by summer of 2021^6^, it is yet unclear how the increasing prevalence of novel SARS-CoV-2 variants^7^, the incidence of break-through infections^8^, and levels of vaccine hesitancy will affect this timeline^9-11^.

Early research around COVID vaccine acceptance has been prospective, with many findings based on surveys about a potential vaccine^12-15^, and/or limited to a population of healthcare workers^16,17^. Recent work has targeted the general population to investigate associations between demographics and attitudes towards the vaccine. Evidence from the United Kingdom (UK) suggests that vaccine hesitancy is higher among younger, lower income, lower education, and non-White minority ethnic groups^18^. The same survey also suggested associations with perceived risk from COVID, reliance on information obtained via specific social media platforms, and levels of trust in government and public health officials. Latent psychological factors, such as religiosity, social dominance, cognitive reflection, and altruism have also been associated with COVID vaccine attitudes in the UK and Ireland^19^.

Besides population characteristics, there is strong evidence that political ideology influences vaccine attitudes. Survey responses from France indicate that the odds of acceptance were highest when respondents identified with the governing party^20^. Recent polls^21,22^ from the US also found stark differences by political affiliation: Democrats – the current governing party – as well as individuals who identified as “liberal” reported being more accepting than Republicans and individuals who identified as “conservative”. However, differences by race/ethnicity (White vs. Black) were comparatively small relative to differences by political ideology and data on racial/ethnic differences from the UK^23^. The extent of racial/ethnic disparities in the US is yet unknown because demographics are available for only 55% of Americans who have received at least one dose. The majority of this group is comprised of White (64%) individuals, followed by Hispanic (12%), Black (9%), and Asian (5%)^24^. Further, documented experimentation at the hands of researchers in communities of color could fuel vaccine hesitancy in the US^25^.

The recent US poll^21^ found that political affiliation is additionally responsible for the variable levels of trust in health experts, broadcast news media, and elected officials with respect to vaccine information. The lack of consensus around credible sources has fueled misinformation around the vaccine, including false claims that vaccines: are linked to cellular networks, are a vehicle to implant microchips, are able to modify the human genome, or that the development process was rushed^26^. In the case of Janssen (Johnson & Johnson), religiosity may influence vaccine uptake given its designation as “morally compromised”^27^. Not surprisingly, exposure to misinformation is harmful to acceptance: an experiment conducted on UK and US participants showed that misinformation reduced intent to get vaccinated; however, women, ethnic minorities, and low-income participants exhibited greater resistance^28^. Additionally, misinformation contributes to fear around vaccination, which in-turn reduces willingness to be vaccinated^22^.

Although US vaccination figures are available through several reliable sources (e.g., CDC^29^, Johns Hopkins Coronavirus Resource Center^30^), there has been a dearth of studies at the sub-state level. Here, we focus on disparities in vaccination rates among US counties. We investigate geographic disparities in average vaccination rates, as well as disparities in the trends of uptake over time, which we call vaccination trajectories. We partition geographic disparities into “expected”, which are attributable to socio-demographic and disease risk factors (e.g., age, educational attainment, COVID incidence rates), and “latent”, which are a function of unobserved or unobservable factors (e.g., political affiliation, levels of trust in officials). In doing so, we identify counties that are over- and under-performing based on their known population characteristics. The estimated vaccination trajectories are then used to forecast when vaccination rates for representative counties are expected to reach a herd immunity threshold of 70%. Consensus around herd immunity for COVID has not yet been established; however, the threshold used in this article is among the plausible levels cited by the Mayo Clinic^31^ and the Kaiser Foundation^32^. Finally, we demonstrate that our predictive model is accurate up to an 8-week-ahead forecast, making our predictions useful for modifying within-state vaccine distribution, adjusting vaccination centers’ staffing needs, and informing investments in local interventions.

## METHODS

### Data sources

Vaccination data were downloaded from the COVID Act Now Application Programming Interface^33^, using an access key obtained in January 2021. COVID Act Now is a nonprofit organization that aggregates data reported by the US Department of Health and Human Services, CDC, and online dashboards for individual states and counties. Data were available for all 50 US states and the District of Columbia, which comprised 3138 (99.9%) counties covering 98.7% of the 2018 US population. Daily vaccinations for each county were aggregated into calendar weeks to avoid day-of-week effects and to reduce noise due to reporting delays and data revisions that may take place within the week. Because county-level data for all 50 states became available only on May 1, our data span the most recent 13 weeks from February 21^st^ through May 16^th^.

Growing evidence points to significant protection from SARS-CoV-2 after one of two planned doses of Pfizer-BioNTech and Moderna^34^, and one planned dose of Janssen. Thus, our outcome variable, was the percentage of county residents with at least 1 dose of these three vaccines, which have emergency use authorization from the US Food and Drug Administration at the time of writing.

### County covariates

Time-invariant demographics were obtained from the 2015 American Community Survey accessed via the choroplethr R package^35^, which included percentages of Hispanic, Black, and Asian residents, and the US Department of Agriculture 2015 Rural Atlas^36^, which included population density, percentages of residents with at most a high school diploma and with at least a 4-year college degree, as well as percentage of Native American residents. Percentage of county residents under age 18 and over age 65 in 2018 was obtained from the CDC Social Vulnerability Index (SVI) webpage^37^, which also contains the estimated 2018 county population, and county polygon boundaries and percentile rankings for the four SVI themes: Socioeconomic Status, Household Composition and Disability, Minority Status and Language, and Housing Type and Transportation. The CDC SVI reflects the socioeconomic conditions of each county and is constructed using 15 county variables, which are current as of 2018. Data were merged using the unique county FIPS code.

Time-varying metrics on COVID incidence rates per 100,000 residents, estimated infection rates (R_t_), and test positivity rates, were obtained from the COVID Act Now API and aggregated into calendar weeks to match the temporal scale of analysis. Mortality rates were available but were excluded due to high correlation with incidence rates.

### Statistical analysis

We used approximate Bayesian models, implemented via Integrated Nested Laplace Approximation (INLA)^38^ in R 4.0^39^ for all estimation. INLA is a popular computing platform in public health for data collected across non-overlapping discrete regions^40-43^. Our models fall within Latent Gaussian Models – the broad class of models for which INLA was developed. Our outcome variable, *Y*_*ct*_, is defined as the proportion of residents for each county *c* (*c* = 1, … *N*_*c*_) at the end of week *t* (*t* = 1, … *N*_*t*_) with at least one dose of the COVID vaccine. A Beta regression framework^44^ was employed, such that *Y*_*ct*_∼*Beta*(*µ*_*ct*_, *ϕ*) with a space-time-varying mean^45,46^ denoted by *µ*_*ct*_

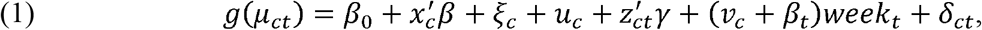

where *g*() is the probit link function and *ϕ* is the Beta regression scale parameter.

In equation (1), *β* are the coefficients of time-fixed covariates, and *ϒ* are the coefficients of time-varying covariates. Spatially structured and unstructured latent county effects are captured by *ξ*_*c*_ and *u*_*c*_, respectively. The former is specified using correlation structures from the Conditionally Autoregressive (CAR)^47^ class with the county adjacency matrix as an input; the latter is specified as an exchangeable random intercept. Jointly, these latent effects measure how vaccination rates vary around the expected value under the model; their sum measures whether each county is over-performing (*ξ*_*c*_ + *u*_*c*_ > 0), or under-performing (*ξ*_*c*_ + *u*_*c*_ < 0),) expectations based on known characteristics.

The coefficient *β*_*t*_ measures the trend in vaccination rates over time – the shape of the vaccination trajectory – for the typical county. Random “slopes” *ν*_*c*_ modify *β*_*t*_ such that each county is allowed its own trajectory (*ν*_*c*_ + *β*_*t*_). Counties where *ν*_*c*_ > 0 have more favorable (i.e., steeper) trajectories than the typical county and vice-versa. The term *δ*_*ct*_ captures space-time (county-week) variability in mean vaccination rates and is specified as a random intercept.

We computed observed and forecasted trajectories for representative counties that fall into different percentiles of the trajectory distribution: top 10%, top 25%, median, bottom 25%, and bottom 10%. Each percentile threshold captures several counties; we selected a representative county such that the posterior SD of its random slope (*ν*_*c*_) is smallest. Once the representative county is selected (*c* = *c*^*^), we construct its trajectory using its latent parameter estimates for 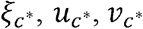 global fixed effects *β*_0_ and *β*_*t*_, assuming all other covariates are held at their average values and averaging over *δ*_*ct*_.

We evaluated several space-time models and selected one using well-established metrics: WAIC and the logarithmic score (LS). We provide more details of our statistical modeling in Supplementary Materials, including: an overview of INLA estimation, prior distributions used, and results from a predictive cross-validation exercise with up to an 8-week-ahead forecast. Diagnostics and parameter estimates for the selected model are shown as part of Supplementary Materials. Additionally, we provide an interactive version of all maps in this article at the following publicly accessible repository (https://github.com/pchernya/Covid_Vaccine_US) that will be updated each week, along with our projections, as new data become available.

## RESULTS

Data from more counties became available over time (Figure 1; Supplementary Table 4); our final counties × weeks sample size was 33055. County covariates were generally uncorrelated: the largest correlations were between percentages of residents under age 18 and over age 65 (*ρ* = −0.58) and percentages of residents with at most a high school diploma and at least a college degree (*ρ* = −0.61) (Supplementary Figure 1); these covariates remained in the statistical model together.

**Figure 1.**
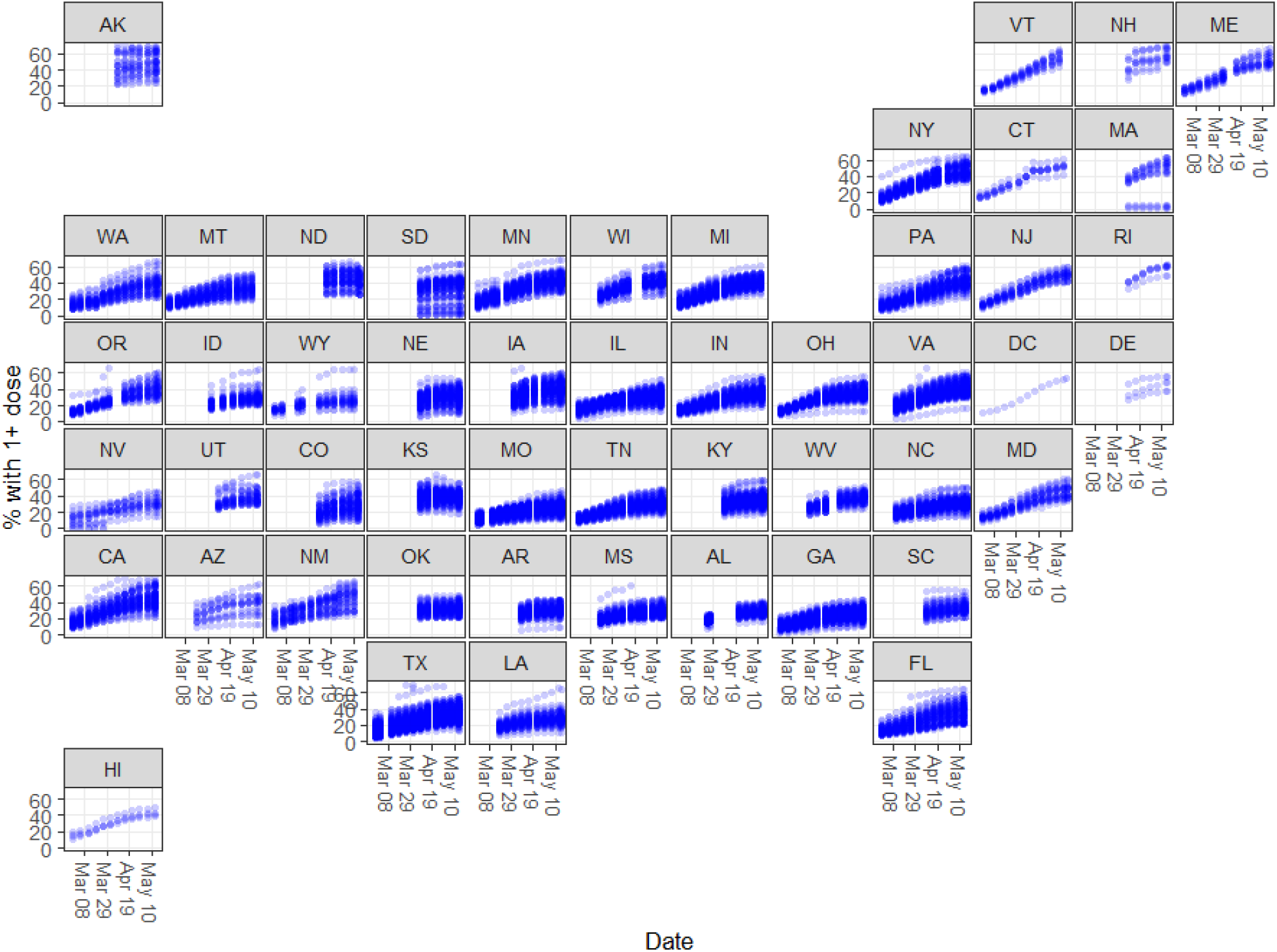
Cumulative percentage of residents with at least one dose of the COVID-19 vaccine by county (N_c_ = 3138) and week (N_t_ = 13), February 21, 2021 through May 16, 2021. Each panel is labelled using a two-letter US state abbreviation. Each point plotted represents data for a county at the end of a given week. Some overplotting may occur; this may lead to an appearance of darker colored points.

### Time-invariant and time-varying coefficients

Counties with high vaccination rates tended to be older, college-educated communities, with high proportions of minority residents, especially Hispanic and Native American individuals (Figure 2). Alternatively, counties with low vaccination rates tended to have low educational attainment and high proportions of children under age 18. Among the time-varying metrics, counties with higher COVID incidence and higher infection rates tended to have higher vaccination rates, whereas counties with high test positivity rates tended to have lower rates.

**Figure 2.**
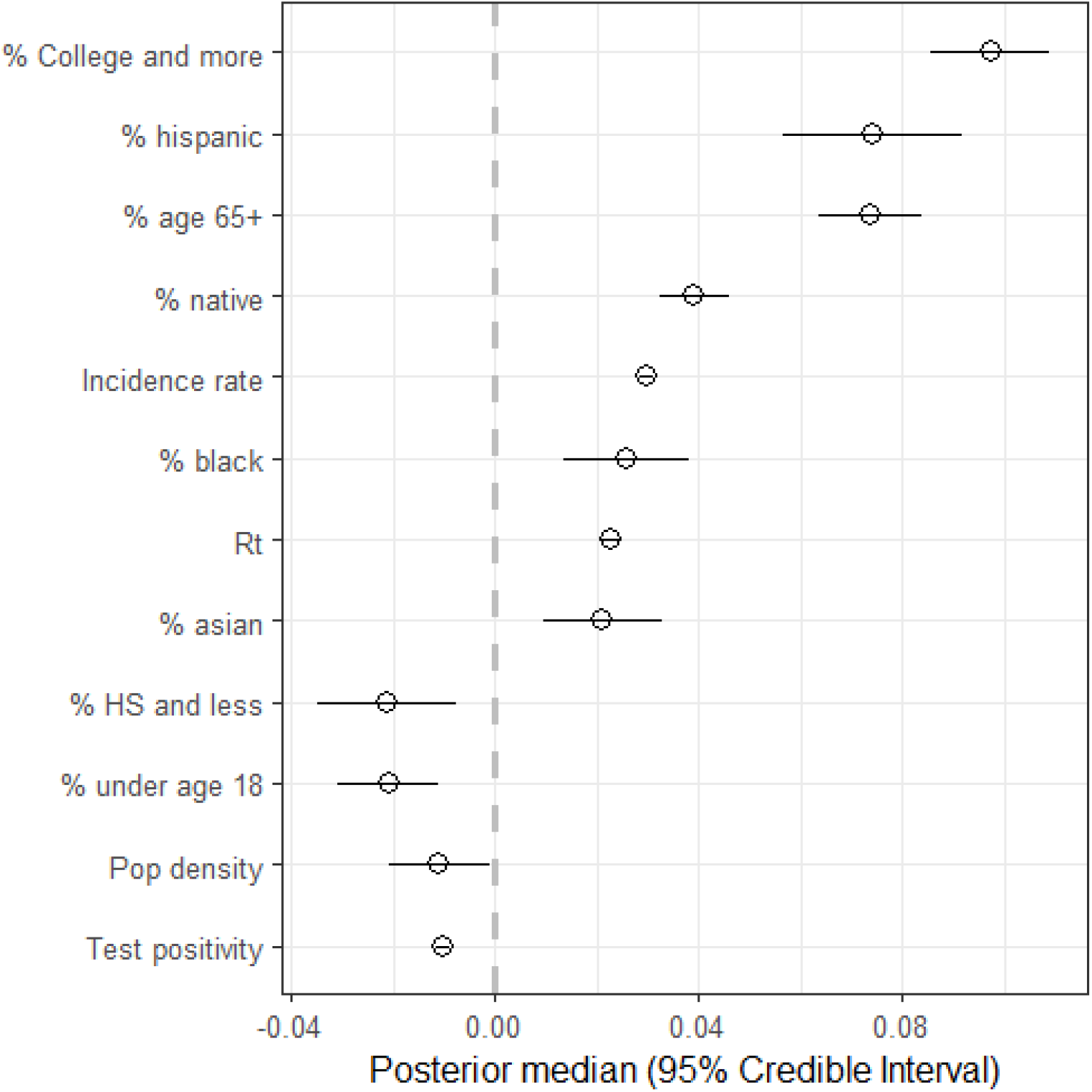
Estimated coefficients of time-fixed and time-varying US county effects from the selected spatio-temporal Beta regression model with a probit link, sorted by absolute magnitude. All covariates were centered and scaled prior to analysis. Vaccination data cover February 21, 2021 through May 16, 2021.

### Latent county effects

Latent county effects and vaccination trajectories reveal strong regional patterns. The majority of counties in New England, Middle Atlantic, and East-North Central US are over-performing, or performing in-line with their known characteristics (Figure 3A). With the exception of Arkansas, Oklahoma, and South Carolina, counties in South Atlantic, East, and West South Central are under-performing their characteristics. Evidence from western US is mixed: counties in California, Kansas, New Mexico, and Utah are largely over-performing, counties in Arizona, Colorado, Nevada, and Wyoming are largely underperforming, and the remaining states have mixed localized patterns. For example, Nebraska, North Dakota, and South Dakota have strong East-West gradients where counties in the East are over-performing expectations and vice-versa.

**Figure 3.**
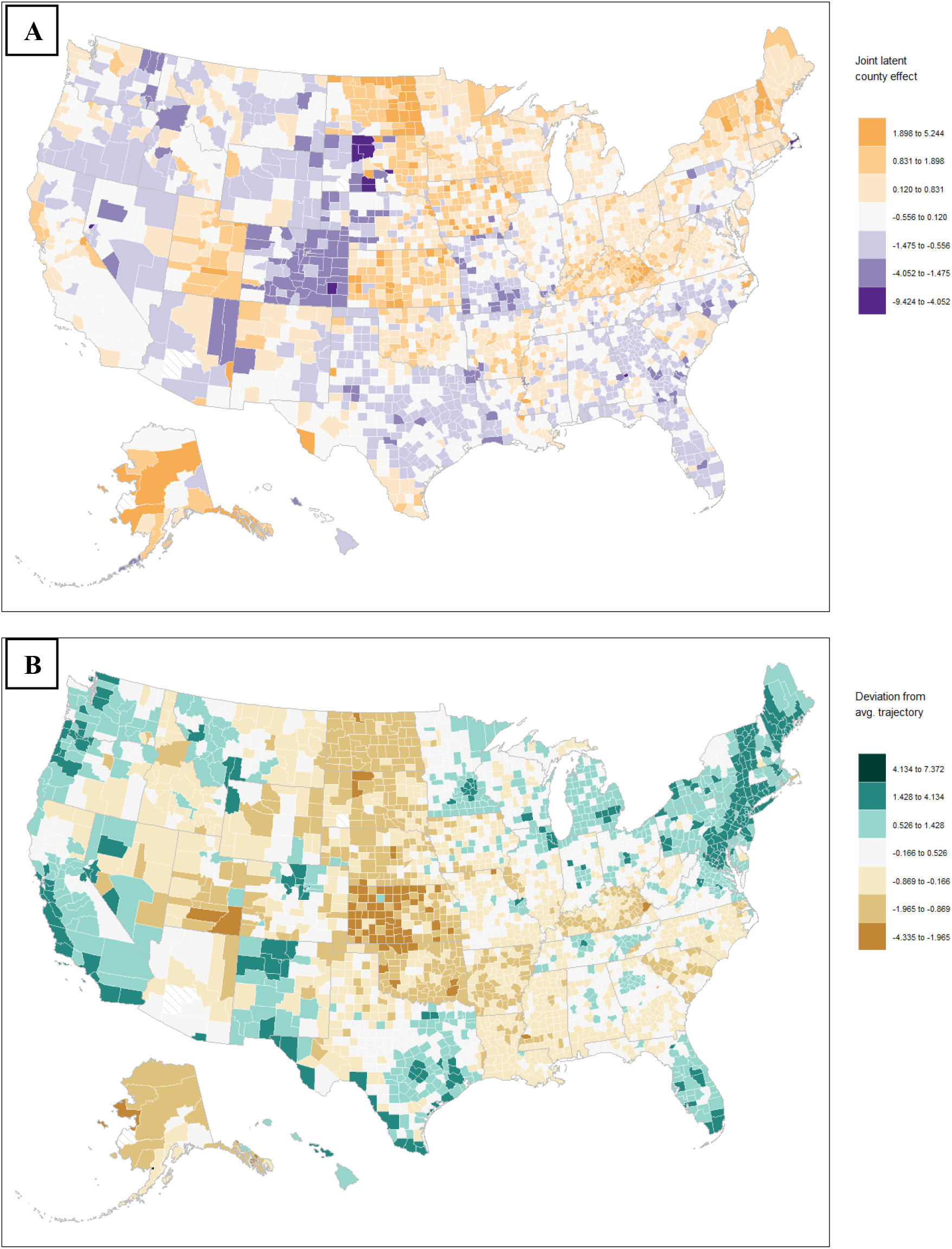
Standardized time-averaged joint (spatial + unstructured) county effects (A) and standardized deviations from the typical trajectory (B). Values > 0 indicate a county is over-performing its known characteristics; values < 0 indicate a county is under-performing its known characteristics. Diagonal hatch pattern indicates missing data, which occurs in Alaska, Arizona, and South Dakota. Vaccination data cover February 21, 2021 through May 16, 2021.

Among the 10 most over-performing counties, 7 are sparsely populated communities in Alaska and North Dakota, and the remaining 3 are in Iowa, New Hampshire, and Virginia (Table 1). The CDC SVI suggests that communities in Alaska face challenges related to housing and transportation and minority and language status. Both counties in North Dakota have favorable rankings across the four SVI themes. The county in Virginia stands out due to its minority and language status, whereas the county in New Hampshire stands out due to its household composition and disability status, as well as its housing and transportation status.

**Table 1.** Top 10 counties according to the time-averaged joint (spatial + unstructured) latent effects, as of May 16, 2021. Latent effects measure over- or under-performance under the regression model; counties in this table have overperformed their known characteristics under the model by at least 3SD. County population (2018) and the 2018 CDC Social Vulnerability Index (SVI) percentiles for Themes 1 through 4 are provided for context. SVI percentiles near 0 indicate lowest social vulnerability; percentiles near 1 indicate greatest social vulnerability.

| Rank | State | County name | Population | Theme 1<br>(Socioeconomic<br>Status) | Theme 2<br>(Household<br>composition<br>& Disability) | Theme 3<br>(Minority<br>status &<br>language) | Theme 4<br>(Housing &<br>transportation) |
| --- | --- | --- | --- | --- | --- | --- | --- |
| 1 | VA | Manassas Park City | 16423 | 0.461 | 0.349 | 0.978 | 0.261 |
| 2 | IA | Cass | 13191 | 0.270 | 0.428 | 0.300 | 0.180 |
| 3 | AK | Skagway | 1061 | 0.153 | 0.010 | 0.392 | 0.812 |
| 4 | AK | Yakutat City | 689 | 0.212 | 0.406 | 0.766 | 0.961 |
| 5 | NH | Coos | 32038 | 0.450 | 0.679 | 0.330 | 0.698 |
| 6 | AK | Petersburg Borough | 3255 | 0.164 | 0.893 | 0.766 | 0.930 |
| 7 | ND | Dickey | 4970 | 0.081 | 0.160 | 0.152 | 0.444 |
| 8 | AK | Sitka City | 8738 | 0.058 | 0.192 | 0.727 | 0.942 |
| 9 | AK | Nome | 9925 | 0.896 | 0.539 | 0.883 | 0.910 |
| 10 | ND | Cavalier | 3824 | 0.048 | 0.119 | 0.251 | 0.037 |

Vaccination trajectories tended to be more favorable in New England, Middle Atlantic, along the Pacific coast, and also in the majority of Florida, Hawaii, Michigan, Minnesota, and New Mexico (Figure 3B). Counties that cover populous metropolitan areas have more favorable trajectories than the surrounding counties, for example: Atlanta, GA; Austin, TX; Chicago, IL; Columbus, OH; Dallas, TX; Denver, CO; Houston, TX; Nashville, TN; and, St. Louis, MO.

Except for counties in Alaska, Nevada, and New Mexico, the 10 most favorable trajectories are generally in the Northeast (Table 2). These counties tend to have favorable socioeconomic status, household composition and disability rankings, as well as housing and transportation rankings, with notable exceptions in Alaska and Vermont. Manassas Park City in Virginia stands out because it is also the most over-performing county in Table 1.

**Table 2.**
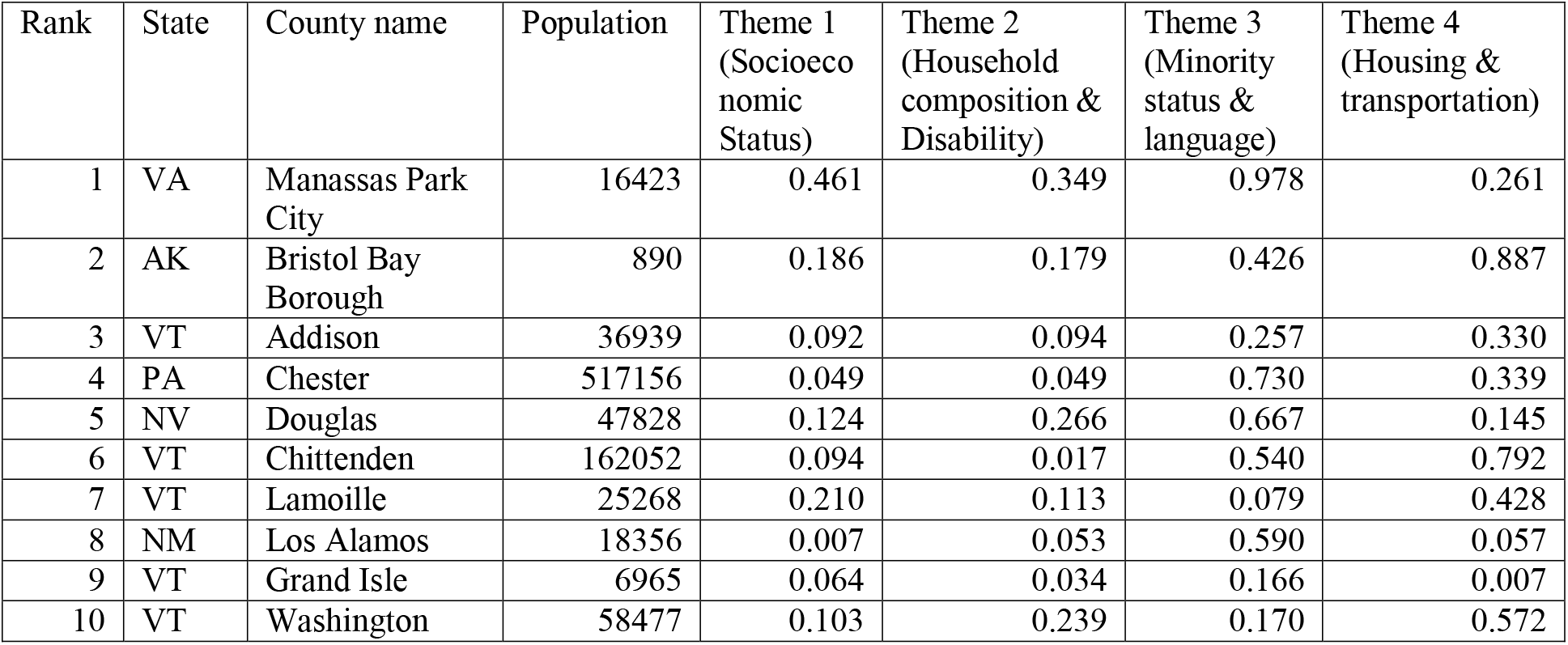
Top 10 counties with most favorable vaccination trajectories, as of May 16, 2021. Counties in this table have the largest positive deviations above the typical vaccination trajectory, by at least 2.8SD. Population (2018) and the 2018 CDC Social Vulnerability Index (SVI) percentiles for Themes 1 through 4 are provided for context. SVI percentiles near 0 indicate lowest social vulnerability; percentiles near 1 indicate greatest social vulnerability.

| Rank | State | County name | Population | Theme 1<br>(Socioeconomic Status) | Theme 2<br>(Household composition & Disability) | Theme 3<br>(Minority status & language) | Theme 4<br>(Housing & transportation) |
| --- | --- | --- | --- | --- | --- | --- | --- |
| 1 | VA | Manassas Park City | 16423 | 0.461 | 0.349 | 0.978 | 0.261 |
| 2 | AK | Bristol Bay Borough | 890 | 0.186 | 0.179 | 0.426 | 0.887 |
| 3 | VT | Addison | 36939 | 0.092 | 0.094 | 0.257 | 0.330 |
| 4 | PA | Chester | 517156 | 0.049 | 0.049 | 0.730 | 0.339 |
| 5 | NV | Douglas | 47828 | 0.124 | 0.266 | 0.667 | 0.145 |
| 6 | VT | Chittenden | 162052 | 0.094 | 0.017 | 0.540 | 0.792 |
| 7 | VT | Lamoille | 25268 | 0.210 | 0.113 | 0.079 | 0.428 |
| 8 | NM | Los Alamos | 18356 | 0.007 | 0.053 | 0.590 | 0.057 |
| 9 | VT | Grand Isle | 6965 | 0.064 | 0.034 | 0.166 | 0.007 |
| 10 | VT | Washington | 58477 | 0.103 | 0.239 | 0.170 | 0.572 |

### Forecasted trajectories

To illustrate the implications of the disparity in vaccination trajectories, we forecasted vaccination rates for counties in the top 10%, top 25%, median, bottom 25%, and bottom 10% of the estimated trajectories distribution (Figure 4). Using 70% as the herd immunity threshold, counties in the top 10% and top 25% are forecasted to reach herd immunity by the week of June 13-19 (Figure 4B I) and July 25-31 (Figure 4B II), respectively. Counties with median trajectories are expected to reach herd immunity by August 8-14 (Figure 4B III), but counties in the bottom 25% of estimated trajectories are only expected to clear the 40% threshold at that time. If current trends hold, these bottom-25% counties are not expected to reach herd immunity until the last week of 2021 (Figure 4B IV), whereas counties in the bottom 10% take until later in 2022.

**Figure 4.**
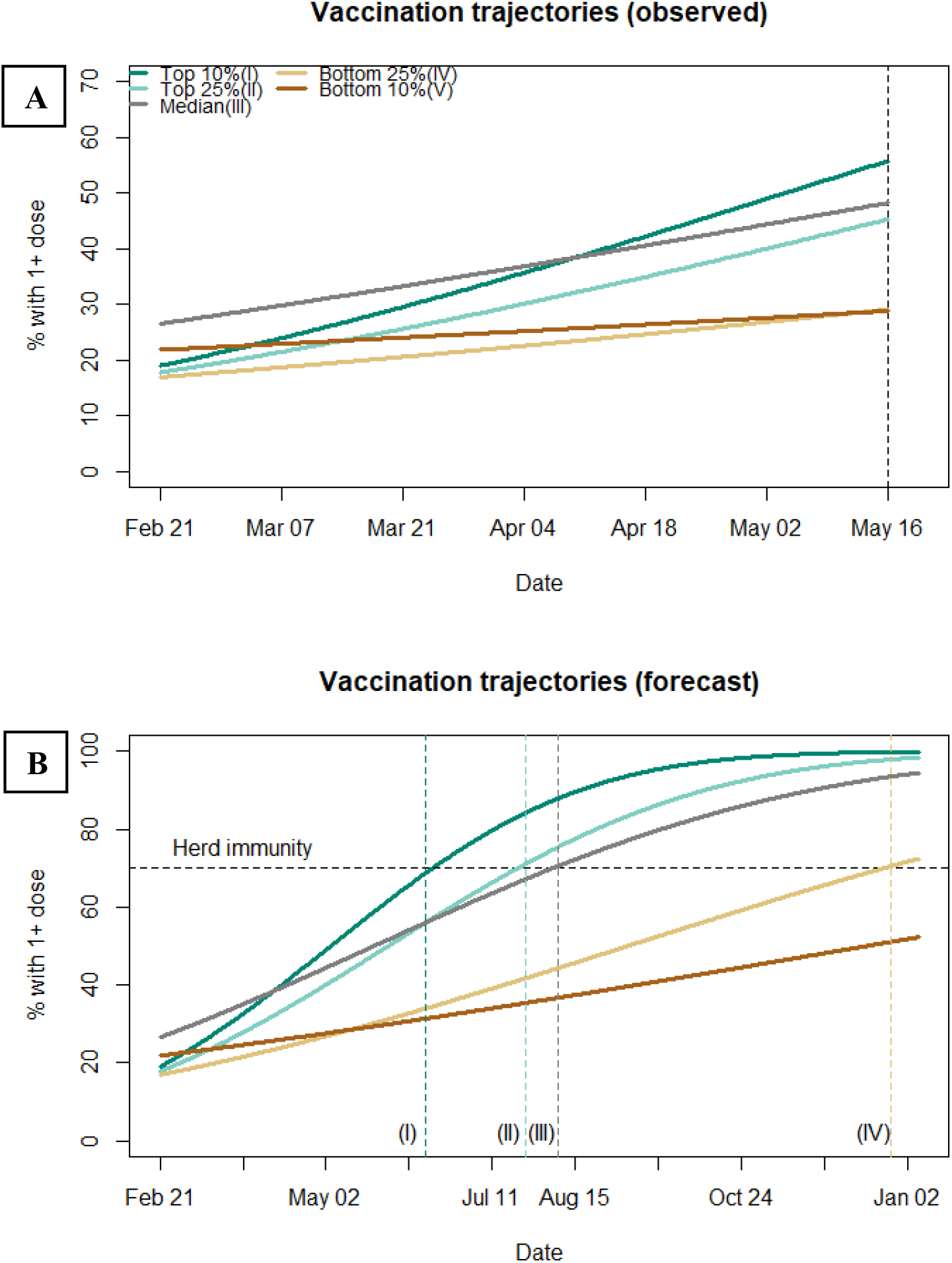
Fitted and forecasted trajectories for representative counties in the top 10%, top 25%, median, bottom 25%, and bottom 10% of estimated trajectories. Vaccination rates for the previous 13 weeks of data as of May 16, 2021 (A) and forecasted vaccination rates (B) are shown. Dashed horizontal line indicates the herd immunity threshold of 70%. Counties in the top 10% of estimated trajectories are expected to reach herd immunity by the week of Jun 13–19 (I); top 25% by the week of Jul 25–31 (II); median county by the week of Aug 8–14 (III); bottom 25% by the week of Dec 26,2021 – Jan 1, 2022 (IV); bottom 10% not until later in 2022.

The forecast for the end of July 2021 (Figure 5A) shows that modal vaccination rates are expected to be between 40% and 60%, with some areas as low as 20%. The herd immunity forecast (Figure 5B) indicates that local immunity can be expected for: most of the counties in the Northeast, coastal Pacific counties, coastal Florida counties, the majority of southern border counties in Texas and New Mexico, and the majority of Hawaii. Additionally, urban areas of Colorado, Illinois, Indiana, Michigan, Minnesota, Ohio, Tennessee, and Texas are also expected to attain local herd immunity.

**Figure 5.**
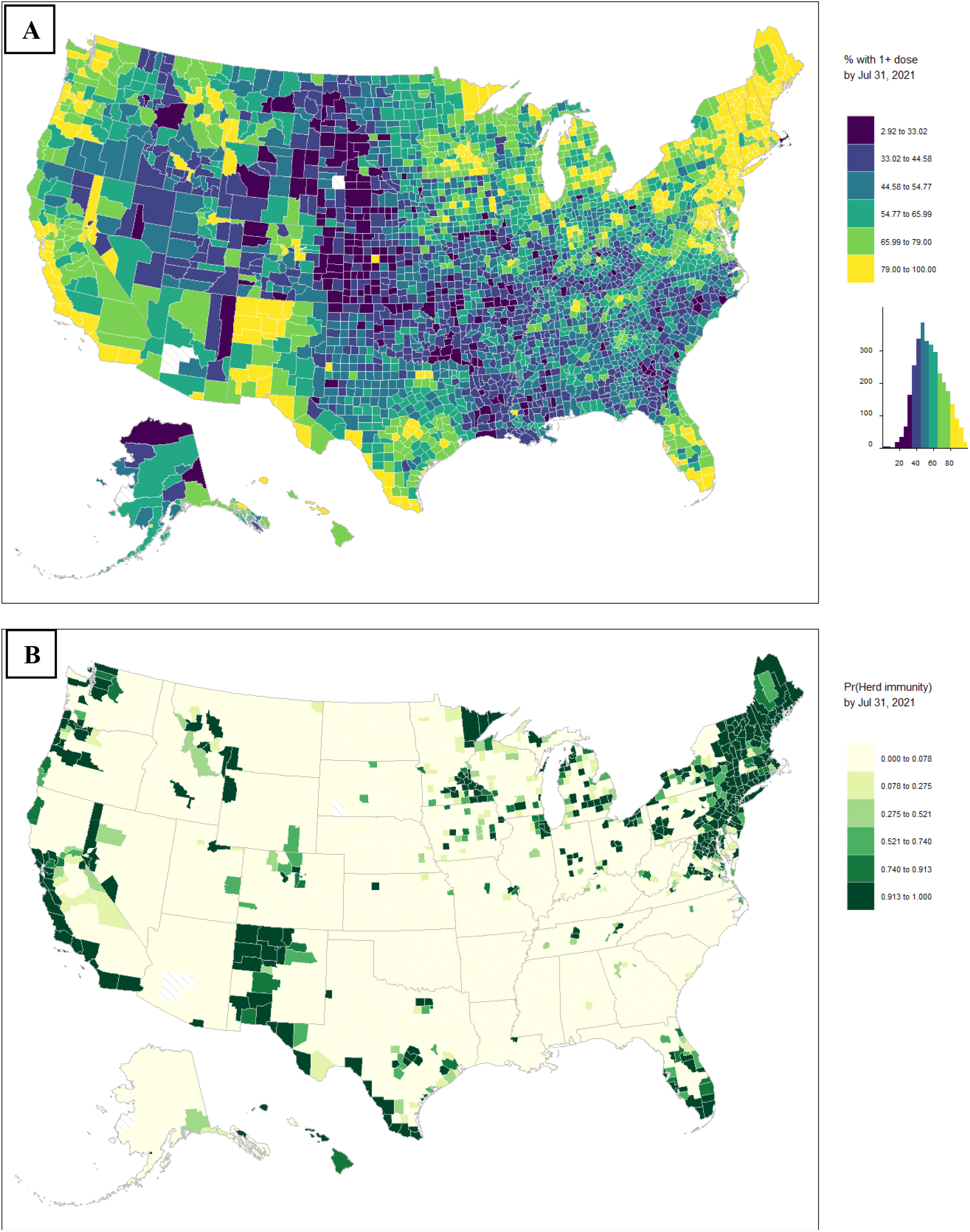
Forecasted percentage of residents with at least one dose by US county for the week ending on July 31 (July 25 – 31) (A) and estimated probability of local herd immunity, i.e. Pr(prediction > 70%) (B). Diagonal hatch pattern indicates missing data, which occurs in Alaska, Arizona, and South Dakota. Forecast is based on data from February 21, 2021 through May 16, 2021.

Our model was accurate for 1-, 2-, and 4-week ahead forecasts, with mean absolute predictive errors of 3.82%, 5.98%, and 9.45% and predictive Pearson correlations of 0.98, 0.97, and 0.95, respectively (Supplementary Table 2; Supplementary Figure 2). Forecast accuracy deteriorated for the 8-week ahead forecast, with mean absolute predictive error of 16.05% and predictive Pearson correlation of 0.64.

## DISCUSSION

To our knowledge, our article describes the first predictive spatio-temporal analysis of US county COVID vaccination uptake. We identified demographic factors and disease risk metrics that explain variability in county vaccination rates and determined counties that over- and under-perform their demographics due to unobserved local factors. Geographic disparities in average vaccination rates and vaccination trajectories were stark, with the latter revealing localized favorable trends concentrated in metropolitan areas. Our forecasts show a growing divide between high-uptake and low-uptake counties: those in the top 25% of estimated vaccination trajectories are expected to reach a 70% herd immunity threshold by the end of July 2021, but those in bottom 25% only by the first week of 2022. Although the median trajectory counties are expected to reach herd immunity by the start of the Fall 2021 school year, these counties are generally concentrated in only a few areas of the country and so the next school year may be adversely affected across a large portion of the US.

Intuitively and in-line with UK data^18^, we show that older and more educated communities have had higher vaccine uptake to date, whereas younger and less educated communities have had lower uptake. This is not surprising given that vaccinations first available to older adults and the use of online sign-up systems are barriers to low-income and communities of color^48,49^. Additionally, county vaccination rates change with COVID risk levels in the community as counties with higher incidence and infection rates have higher vaccination rates, on average. This suggests that greater visibility of current community risk via local media channels may be beneficial.

Less intuitively, we offer evidence that counties with large minority populations, especially Hispanic, Native, and Black residents, have higher vaccination uptake. This result might be attributed to health officials’ efforts to reach minority populations early with targeted messaging^50^ plus the fact that minorities have had a disproportionate number of cases and deaths during the pandemic. Consistent with greater vaccination rates among residents who perceive greater risk^19^, minority communities may be more accepting of the COVID vaccine than suggested by prior surveys^21^.

Our finding of negative association between test positivity and vaccination rates may also be unintuitive. In our view, this variable acts as a proxy for testing behavior as opposed to a measure of community risk. Counties with high test positivity may signal that it is difficult to obtain a test, or residents tend to get tested only after symptoms have developed. This may also reflect the perception that a positive test indicates one will become immune after several weeks and so residents are less likely to obtain a vaccination.

Our ability to score each county as over- or under-performing their known characteristics can inform vaccine redistribution efforts within- and between-states. Our models were accurate within a 4-weeks-ahead forecast and may still be useful up to an 8-week-ahead forecast. Predicted vaccination rates will be updated on a weekly basis on this website (https://github.com/pchernya/Covid_Vaccine_US). In addition to vaccine redistribution, these can inform local reopening policies, and staffing levels at vaccination sites.

Our article has several important limitations. First, our data are ecological in nature, only available at the level of county. Thus, we cannot use our analysis to infer behavior for any individual resident. Second, while the CDC reports making reasonable efforts to determine county of residence, certain reporting practices, such as reporting through retail pharmacies, federal facilities, or long-term care facilities may result in low vaccination coverage data. This has potential to add noise to our vaccination maps, especially in areas of high resident mobility or vaccination tourism. Third, although we produce accurate short-term forecasts, unforeseen factors such as discoveries of major adverse reactions, geopolitical events, natural disasters, and logistical breaks in the supply chain cannot be incorporated into our forecast, although these will certainly affect vaccination uptake. In fact, our longer-term forecast should only be considered as a counter-factual scenario if current trends hold.

## Data Availability

Data can be made available for research purposes after publication upon reasonable request from the corresponding author.

https://github.com/pchernya/Covid_Vaccine_US

## Notes

### Competing Interest Statement

The authors have declared no competing interest.

### Funding Statement

None to report.

### Author Declarations

No IRB/oversight body information to report.

