## Supplementary Materials for "COVID-19 vaccine uptake in United States counties: geospatial vaccination patterns and trajectories towards herd immunity"

**Supplementary Methods**

*Model specification*

Our data are spatio-temporal, with counties serving as spatial components $\left( c=1,\ldots,N_{c} \right)$ and weeks serving as temporal components $\left( t=1,\ldots,N_{t} \right)$. The outcome variable is the proportion of residents in county $c$ at the end of week $t$ with at least one dose of the vaccine ($Y_{ct}$), for which we assume a Beta distribution: $Y_{ct}\sim Beta(\mu_{ct},\phi)$. In a Beta regression model, the mean proportion is modelled as $E\left( Y_{ct} \right)=g\left( \mu_{ct} \right)=x_{ct}^{'}\beta$, where the link function $g\left( \cdot\right)$ ensures that the mean remains strictly between 0 and 1, and the variance is modelled as $Var\left( Y_{ct} \right)=\frac{\mu_{ct}(1-\mu_{ct})}{1+\phi}$. Exploratory analyses suggested that the probit link performs better than the logit link, so $g\left( \mu\right)=\Phi(\mu)$, where $\Phi(\cdot)$ is the Cumulative Distribution Function of the standard Normal.

Methods to model spatio-temporal public health outcomes are well developed (e.g., Shrödle and Held, 2011; Blangiardo et al., 2013; Ugarte et al., 2014; Bakka et al., 2018); adapting the specifications described therein, our mean proportion was specified as follows:

(1)	$g\left( \mu_{ct} \right)={\beta_{0}+x}_{c}^{'}\beta+z_{ct}^{'}\gamma+\xi_{c}+u_{c}+\left( v_{c}+\beta_{t} \right)week_{t}+\delta_{ct}$

$$\boldsymbol{\xi}\sim MVN(\boldsymbol{0},\boldsymbol{\Sigma}_{\xi})$$

$$\left[ \begin{matrix} \boldsymbol{u} \\ \boldsymbol{v} \end{matrix} \right]\sim MVN\left( \boldsymbol{0}, \left[ \begin{matrix} \sigma_{u}^{2} & \rho\sigma_{u}\sigma_{v} \\ \rho\sigma_{u}\sigma_{v} & \sigma_{v}^{2} \end{matrix} \right] \right)$$

$$\boldsymbol{\delta}\sim MVN(0, \sigma_{\delta}^{2}\mathbf{I})$$

For each county, the time-invariant and time-varying fixed effects are contained in $x_{c}^{'}$ and $z_{ct}^{'}$, respectively, and $\beta$ and $\gamma$ are the corresponding coefficients. Variability in latent county effects is typically partitioned between spatially structured ($\xi_{c}$) and unstructured effects ($u_{c}$), with the joint county effects represented by $\xi_{c}+u_{c}$. Spatial correlation between adjacent counties can be captured using a Conditionally Auto-regressive (CAR) model, which imply one of the following covariance structures for the joint effects ($\boldsymbol{\Sigma}_{\xi+u}$):

i) Besag-York-Mollié (BYM): $\boldsymbol{\Sigma}_{\xi+u}=\sigma_{\xi}^{2}\left( \mathbf{D}-\mathbf{W} \right)^{-}+\sigma_{u}^{2}\mathbf{I}$
ii) Leroux + exchangeable: $\boldsymbol{\Sigma}_{\xi+u}=\sigma_{\xi}^{2}\left[ \lambda\left( \mathbf{D}-\mathbf{W} \right)^{-}+\left( 1-\lambda\right)\mathbf{I} \right]^{-1}+\sigma_{u}^{2}\mathbf{I}$

In the BYM model (Besag et al., 1991), $\sigma_{\xi}^{2}$ and $\sigma_{u}^{2}$ measure the spatial and unstructured variability, respectively. The proportion of the total variance attributed to spatial structure is $\frac{\sigma_{\xi}^{2}}{\sigma_{\xi}^{2}+\sigma_{u}^{2}}$. In the Leroux CAR model (Leroux et al., 2000), $\sigma_{\xi}^{2}$ measures between-county variability and $\lambda$ is a mixing parameter that reflects the proportion of total variance that may be attributed to the county adjacency-based spatial structure (encoded in the $N_{c}\times N_{c}$ queen-style adjacency matrix **W**).

Parameters $v_{c}$ are the county random slopes, which may be correlated to the unstructured county effects $u_{c}$. The weekly trend ($\beta_{t}$) for the “typical” county is modified by the county-specific effect $v_{c}$, allowing each county to have its own trend over time, and thus its own vaccination trajectory. The term $\delta_{ct}$ captures latent spatio-temporal variability across counties and weeks, which is modelled as an exchangeable random effect (e.g., Blangiardo et al., 2013).

In our model selection exercise (Supplementary Table 1), we compare several versions of the general model shown in equation (1): BYM vs. Leroux CAR model, and also models that treat $u_{c}$ and $v_{c}$ as uncorrelated (i.e., $\rho=0$). We present the Watanabe-Akaike Information Criterion (Watanabe, 2013; WAIC), the logarithmic score (Gneiting and Raftery, 2007; LS), the log-marginal likelihood (LML), and the effective number of parameters under the WAIC formulation for each model. The preferred model has the smallest information criteria and LS and is expected to have the smallest out-of-sample predictive error. Finally, we also report computing time in minutes for each model estimated.

*Estimation and inference via Integrated Nested Laplace Approximation (INLA)*

Our models fall into the class of Latent Gaussian Models and so the estimation task lends itself well to approximate Bayesian inference via Integrated Nested Laplace Approximation (INLA; Rue et al., 2009). INLA has been widely used for a variety of spatio-temporal models and is especially popular where non-overlapping spatial areas form a lattice. We note that our model can be estimated using Markov Chain Monte Carlo sampling; however, in our experience with even the most computationally efficient NUTS HMC, computing times are orders of magnitude longer than those using the R-INLA package. We briefly describe the INLA modeling framework here and direct the reader to previously published texts (e.g., Blangiardo and Cameletti, 2015; Lawson, 2021) for a more comprehensive overview.

Let $\boldsymbol{Y}=\left( Y_{11},\ldots,Y_{N_{c}{\times N}_{t}} \right)'$ denote the vector of observed proportions of county residents with at least one dose and let $\boldsymbol{\eta}=\left\{ \boldsymbol{\xi},\boldsymbol{u},\boldsymbol{v},\boldsymbol{\delta} \right\}$be the multivariate latent Gaussian random field. Let the vector $\boldsymbol{\theta}=\left\{ \boldsymbol{\theta}_{1},\boldsymbol{\theta}_{2} \right\}$ contain the hyperparameters that govern the likelihood of $\boldsymbol{Y}$ and the latent Gaussian field $\boldsymbol{\eta}$. Using a hierarchical formulation, the likelihood of $\boldsymbol{Y}$ is assumed to be conditionally independent given the latent field and the hyperparameters:

(2) $\boldsymbol{Y|\eta,}\boldsymbol{\theta}_{1}\sim\prod_{i} f\left( Y_{i}|\eta_{i},\boldsymbol{\theta}_{1} \right), i=1,\ldots,N_{c}\times N_{t}$

In our application, the conditional likelihood is assumed to be the Beta distribution because our outcome is a vector of proportions, and so $\boldsymbol{\theta}_{1}$ consists only of the scale parameter $\phi$. The mean of the Beta distributions is an inverse-linked linear combination of fixed and random effects, as shown in equation (1). The latent field is assumed to be Normally distributed with mean $\boldsymbol{0}$, and the precision matrix $\boldsymbol{Q}\left( \boldsymbol{\theta}_{2} \right)$:

(3) $\boldsymbol{\eta|}\boldsymbol{\theta}_{2}\sim N\left( \boldsymbol{0},\boldsymbol{Q}\left( \boldsymbol{\theta}_{2} \right)^{-1} \right)$

For computational efficiency and to improve accuracy of the Laplace approximation, the dimension of $\boldsymbol{\theta}$ must be small, preferably around 5 and less than 20 (e.g., Bakka, 2018). In our application, using the most general model with the Leroux CAR spatial correlation structure, spatio-temporal interaction $\boldsymbol{\delta}$, and correlation between unstructured spatial effects and random slopes, $\boldsymbol{\theta}$ contains 7 parameters: $\boldsymbol{\theta=}\left\{ \boldsymbol{\theta}_{\boldsymbol{1}}\boldsymbol{,}\boldsymbol{\theta}_{\boldsymbol{2}} \right\}=\left\{ {\phi,\lambda,\sigma}_{\xi}^{-2},\sigma_{u}^{-2},\sigma_{v}^{-2},\rho,\sigma_{\delta}^{-2} \right\}$. To complete the hierarchical formulation, prior distributions are assigned to elements of $\boldsymbol{\theta}$ that reflect prior knowledge about the parameters:

(4) $\boldsymbol{\theta}\sim\pi(\boldsymbol{\theta})$

These priors are typically assumed to be *apriori* independent and were specified here as follows:

- Penalised-complexity (PC) priors (Simpson et al., 2014) for all precision parameters, generically ($\tau$), such that Prob($\frac{1}{\sqrt{\tau}}>1)=0.01$.
- Uninformative logit-normal prior for Leroux mixing parameter lambda: $logit\left( \lambda\right)\sim N(\mu=0, \sigma= 2.5)$
- Weakly-informative priors for the intercept and all fixed effects: $\beta_{0}\sim N(0, 10)$; $\beta_{1}\ldots\beta_{9}\sim N(0, 10)$; $\gamma_{1}\ldots\gamma_{3}\sim N(0, 10)$
- Weakly-informative prior the log-scale parameter $\phi$: $\log\left( \phi\right)\sim N(0, 10)$

Because $\boldsymbol{\eta}$ are assumed to be a Gaussian Markov Random Field (GMRF), the joint posterior distribution of the latent field and the hyperparameters can be expressed as: $\pi\left( \boldsymbol{\eta},\boldsymbol{\theta}|\boldsymbol{Y} \right)\propto\pi\left( \boldsymbol{\theta} \right)\left| \boldsymbol{Q}\left( \boldsymbol{\theta}_{2} \right) \right|^{1/2}exp\left\{ -0.5\boldsymbol{\eta}^{'}\boldsymbol{Q}\left( \boldsymbol{\theta}_{2} \right)\boldsymbol{\eta}+\sum_{i} \log\left( f\left( Y_{i}|\eta_{i},\boldsymbol{\theta}_{1} \right) \right) \right\}$. Unlike estimation via MCMC, which attempts to take high-dimensional samples from $\pi\left( \boldsymbol{\eta},\boldsymbol{\theta}|\boldsymbol{Y} \right)$, quantities of interest for INLA lie in the element-wise marginal posterior distributions of the latent field and the hyperparameters given the data, expressed as $\pi(\eta_{i}\boldsymbol{|Y})$ and $\pi(\theta_{j}\boldsymbol{|Y})$ (e.g., see Martino and Riebler, 2019):

(5) $\pi\left( \eta_{i} | \boldsymbol{Y} \right)\propto\int\pi\left( \eta_{i} | \boldsymbol{\theta,Y} \right)\pi\left( \boldsymbol{\theta|Y} \right)d\boldsymbol{\theta,}i=1,\ldots,N_{c}N_{t}$

$\pi\left( \theta_{j} | \boldsymbol{Y} \right)\propto\int\pi\left( \boldsymbol{\theta}|\boldsymbol{Y} \right)d\boldsymbol{\theta}_{j}, j=1,\ldots,dim(\boldsymbol{\theta})$

The integrals in (5) do not generally have closed form solutions and quantities therein must be approximated. By default, INLA employs Laplace approximation for the two marginal posteriors in (5), denoted as $\tilde{\pi}\left( \theta_{j} | \boldsymbol{Y} \right)$ and $\tilde{\pi}\left( \eta_{i} | \boldsymbol{Y} \right)$, using *K* quadrature points that placed around the posterior mode with corresponding quadrature weights $\Delta_{k}$:

(6) $\tilde{\pi}\left( \eta_{i} | \boldsymbol{Y} \right)=\sum_{k} \pi\left( \eta_{i} | \boldsymbol{\theta}^{k}\boldsymbol{,Y} \right)\tilde{\pi}\left( \boldsymbol{\theta}^{k} | \boldsymbol{Y} \right)\Delta_{k}$

$\tilde{\pi}\left( \theta_{j} | \boldsymbol{Y} \right)=\sum_{k} \tilde{\pi}\left( \theta_{j},\boldsymbol{\theta}_{-j}^{k} | \boldsymbol{Y} \right)\Delta_{k}$

We note that the approximation $\tilde{\pi}\left( \boldsymbol{\theta} | \boldsymbol{Y} \right)$ is included into the approximation for the latent field, giving rise to the “nested” nature of INLA computing framework. Aside from known cases where these approximations fail (e.g., binary-valued multi-level models (Fong et al., 2010; Ferkingstad and Rue, 2015) or in the case of multi-modal posteriors), INLA has been found to be accurate and largely concordant with MCMC results (e.g., Schrödle et al., 2010; Schrödle and Held, 2011).

*Evaluating model predictive performance*

To investigate the predictive performance of our chosen statistical model, we perform a cross-validation exercise using the 13 most recent weeks of data. For the 1-week-ahead forecast, we estimate our model using data from weeks 1 through 12 and evaluate predictions for week 13 using observed data from that week. For the 2-, 4-, and 8-weeks-ahead forecast, we estimate the model using weeks 1 through 11, and 1 through 10, and 1 through 5 respectively, and evaluate predictions using observed data for week 13. We note that the spatial scope of prediction increases over time in our cross-validation analysis. In fact, data on only 2205 counties (70%) were incorporated into the 8-weeks-ahead forecast, although 3065 counties (97.6%) were incorporated into the 4-weeks-ahead forecast, and all 3137 counties were incorporated into the 2-weeks-ahead forecast.

In the INLA R package, prediction is performed as part of estimation. So, we replace the observations we want to predict with NA and re-estimated our selected model. Based on the prediction for the 13^th^ week, we report the Root-Mean Squared Error $RMSE=\sqrt{\sum_{c=1}^{N_{c}} \left( Y_{c13}-\hat{Y}_{c13} \right)^{2}}$, Mean Absolute Error $MAE=\sum_{c=1}^{N_{c}} \left| Y_{c13}-\hat{Y}_{c13} \right|$, as well as the Spearman rank order correlation and the Pearson correlation between observed $Y_{c13}$ and predicted $\hat{Y}_{c13}$ for $c=1,\ldots,N_{c}$ (Supplementary Table 2).

**Supplementary Figures and Tables**

Supplementary Figure 1. Visual correlation matrix for time-invariant and time-varying county explanatory variables used for statistical modeling. Only Pearson correlations that are statistically significant ($\alpha=0.05$) are shown; the diagonal is omitted. Vaccination data cover February 21, 2021 through May 16, 2021.

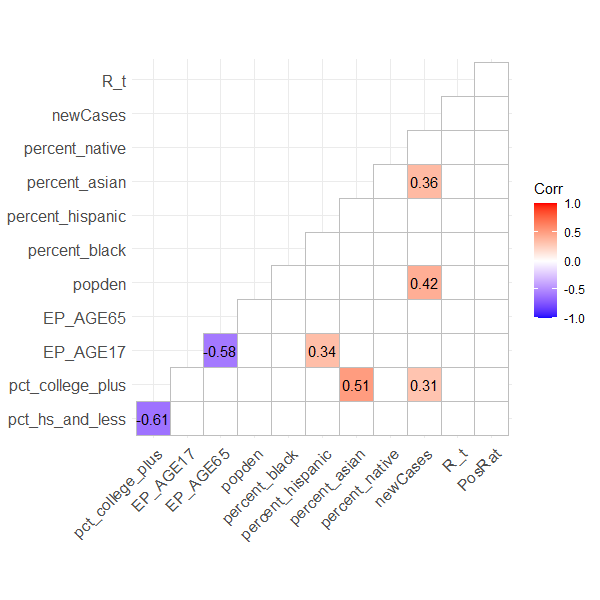

Supplementary Figure 2. Predictive performance of the selected model for the 1-, 2-, 4-, and 8-week-ahead forecast of cumulative proportion of residents with at least one dose. Each forecast targets the 13^th^ week of the dataset. The 1-week forecast is produced using data from all available counties during weeks 1 through 12; 2-week forecast from weeks 1 through 11; 4-week forecast from weeks 1 through 9; and 8-week forecast from weeks 1 through 5. Predictive metrics are reported in Supplementary Table 2. Observed vaccination data cover February 21, 2021 through May 16, 2021.

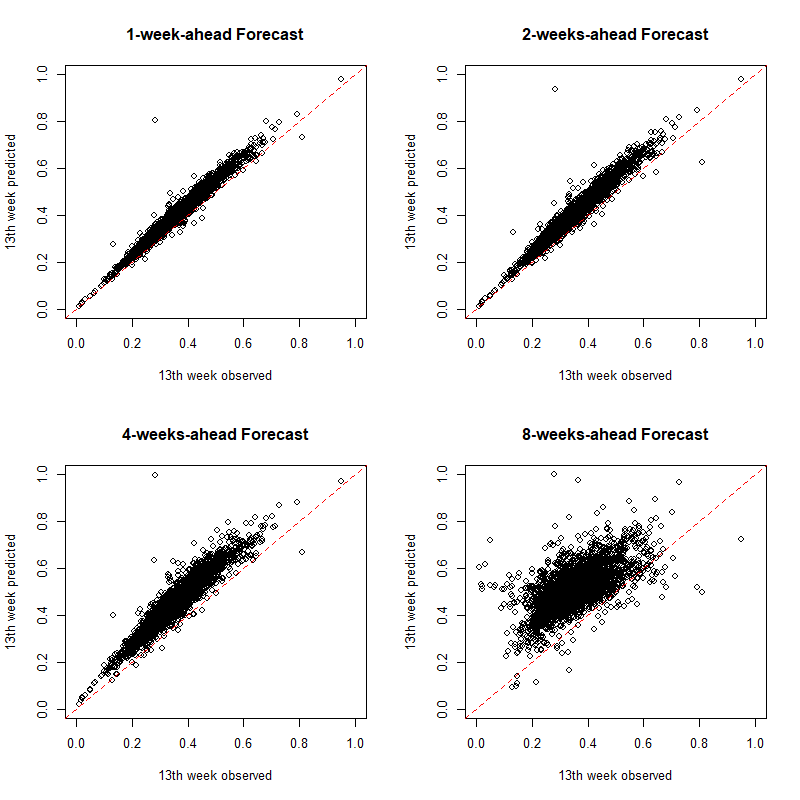

Supplementary Figure 3. COVID-19 incidence rate per week and US state. Incidence is shown on the natural-logarithm scale; each point is a county. Vaccination data cover February 21, 2021 through May 16, 2021.

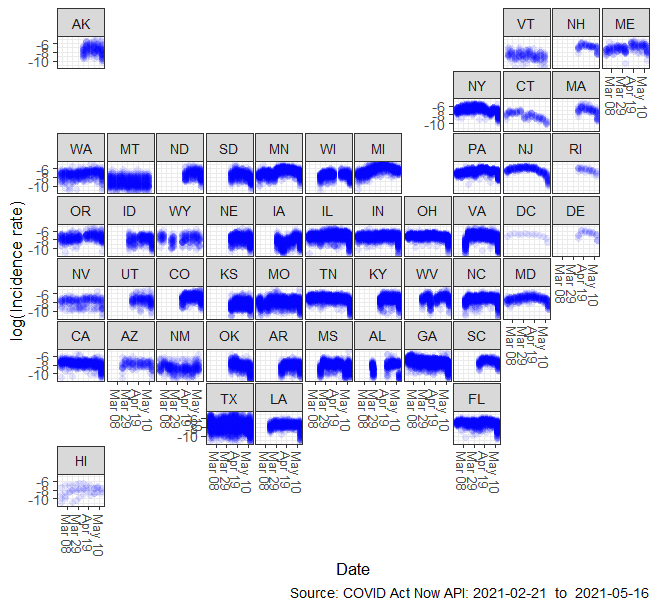

Supplementary Figure 4. COVID-19 test positivity rate per week and US state. Each point is a county. Vaccination data cover February 21, 2021 through May 16, 2021.

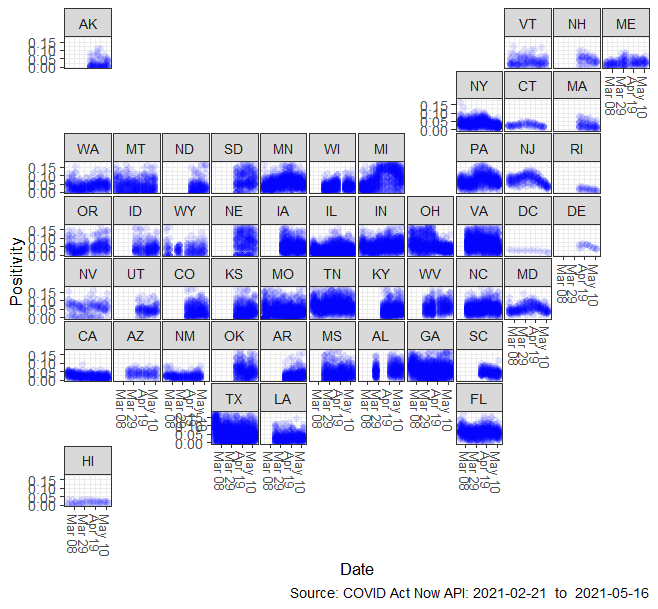

Supplementary Figure 5. COVID-19 estimated infection rate (Rt) per week and US state. Each point is a county. Vaccination data cover February 21, 2021 through May 16, 2021.

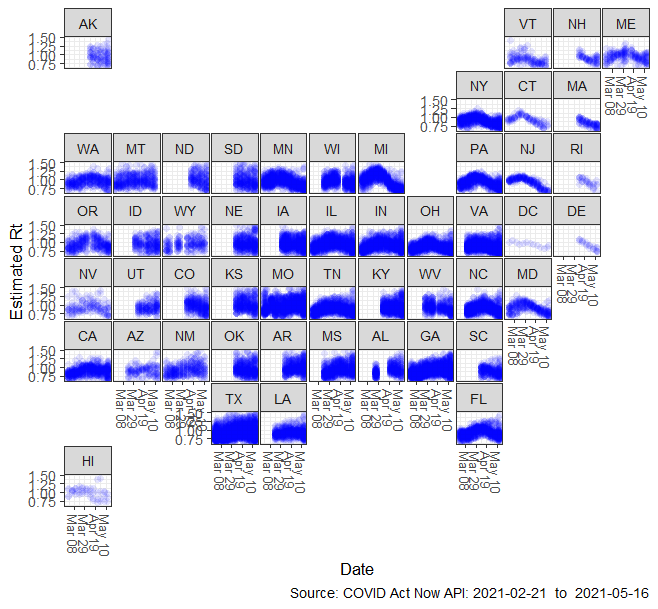

Supplementary Figure 6. Probability Integral Transformation (PIT) to assess model calibration. Except for quantities in the tails, no systematic deviation from a Uniform distribution is detected. Model appears well-calibrated for fitted values between 0 and 0.8. Vaccination data cover February 21, 2021 through May 16, 2021.

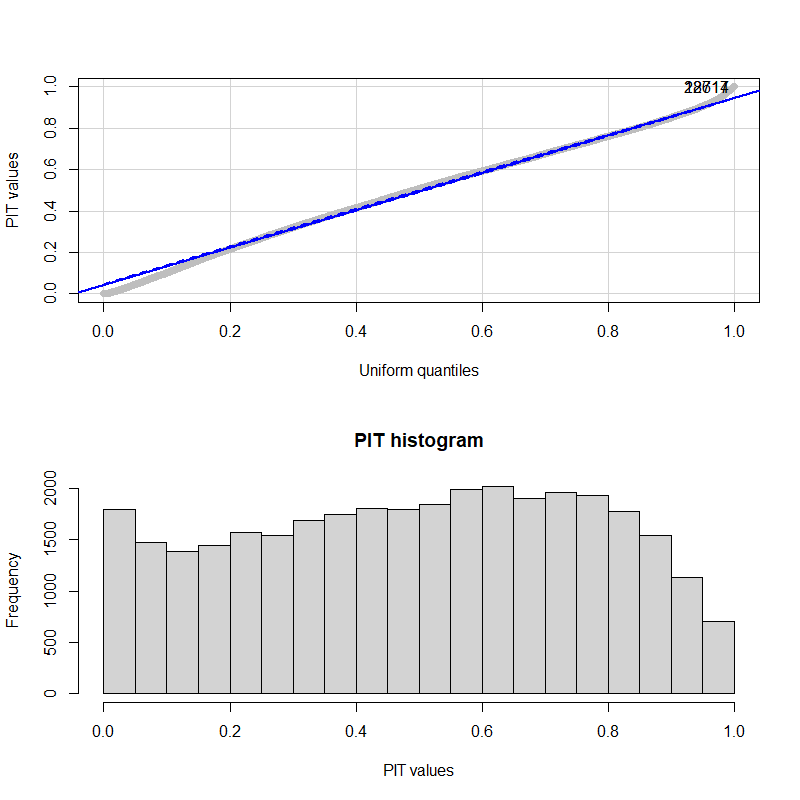

Supplementary Table 1. Model selection metrics for the estimated spatio-temporal models. Models with lower Watanabe-Akaike Information Criteria (WAIC) and logarithmic score (LS) are expected to have lower out-of-sample prediction error and are thus preferred. The column $p_{WAIC}$ denotes the effective number of parameters under the WAIC formulation. LML denotes the log-marginal likelihood via integration. Models with the smaller WAIC and LS are expected to have smaller predictive error and are thus preferred. Vaccination data cover February 21, 2021 through May 16, 2021.

|  | Spatial model | Unstruct Int-Slope Correlation | WAIC | $p_{WAIC}$ | LS | LML | Computing Time (mins) |
| --- | --- | --- | --- | --- | --- | --- | --- |
| **1** | **BYM (ICAR + exch.)** | **No** | **-156652** | **9690** | **-2.346** | **69469.3** | **3.87** |
| 2 | BYM (ICAR + exch.) | Yes | -155297 | 8936 | -2.340 | 66256.2 | 8.93 |
| 3 | Leroux + exch. | No | -155781 | 9640 | -2.346 | 69106.5 | 15.52 |
| 4 | Leroux + exch. | Yes | -156401 | 9449 | -2.345 | 68931.1 | 35.68 |

Supplementary Table 2. Predictive performance of the selected model (**Model 1**) for the 1-week-ahead, 2-weeks-ahead, 4-weeks-ahead, and 8-weeks-ahead forecast. RMSE is the root-mean squared error; MAE is the mean absolute error. The dimension of the spatial and spatio-temporal components increased over time as additional counties reported data. Observed vaccination data cover February 21, 2021 through May 16, 2021.

| Forecast (weeks ahead) | RMSE | MAE | Spearman Rank Corr. | Pearson Corr. |
| --- | --- | --- | --- | --- |
| 1-week | 0.0438 | 0.0382 | 0.9811 | 0.9812 |
| 2-weeks | 0.0598 | 0.0532 | 0.9722 | 0.9704 |
| 4-weeks | 0.1017 | 0.0945 | 0.9496 | 0.9467 |
| 8-weeks | 0.1775 | 0.1605 | 0.6646 | 0.6448 |

Supplementary Table 3. Parameter estimates for the selected model (Model 1): BYM with no correlation between unstructured county intercepts and county random slopes. Parameter estimates reflect results from a Beta regression with a probit link function. All explanatory variables are quantitative and were centered and scaled prior to estimation. Vaccination data cover February 21, 2021 through May 16, 2021. KLD is the Kullback-Leibler divergence between the simplified Laplace and Gaussian approximation for each posterior; values near 0 indicate the approximation was successful.

| Parameter name |  | Posterior median | 95% Credible Interval | KLD |
| --- | --- | --- | --- | --- |
| Intercept | $\beta_{0}$ | -0.5570 | (-0.5618, -0.5522) | 0.00000 |
| Weekly trend | $\beta_{t}$ | 0.1781 | (0.1744, 0.1817) | 0.00000 |
| % HS and less | $\beta_{1}$ | -0.0210 | (-0.0348, -0.0073) | 0.00000 |
| % college and more | $\beta_{2}$ | 0.0974 | (0.0856, 0.1091) | 0.00000 |
| % under age 18 | $\beta_{3}$ | -0.0209 | (-0.0306, -0.0112) | 0.00000 |
| % age 65+ | $\beta_{4}$ | 0.0738 | (0.0636, 0.0841) | 0.00000 |
| Population density | $\beta_{5}$ | -0.0110 | (-0.0209, -0.0011) | 0.00000 |
| % Black | $\beta_{6}$ | 0.0260 | (0.0135, 0.0384) | 0.00000 |
| % Hispanic | $\beta_{7}$ | 0.0743 | (0.0565, 0.0920) | 0.00000 |
| % Asian | $\beta_{8}$ | 0.0212 | (0.0096, 0.0329) | 0.00000 |
| % Native | $\beta_{9}$ | 0.0392 | (0.0324, 0.0459) | 0.00000 |
| Incidence rate per 10^5^ | $\gamma_{1}$ | 0.0300 | (0.0286, 0.0313) | 0.00000 |
| Infection rate (Rt) | $\gamma_{2}$ | 0.0226 | (0.0207, 0.0246) | 0.00000 |
| Positive test % | $\gamma_{3}$ | -0.0100 | (-0.0113, -0.0087) | 0.00000 |
| SD of spatial latent effects | $\sigma_{\xi}$ | 0.2301 | (0.2151, 0.2503) |  |
| SD of unstructured latent effects | $\sigma_{u}$ | 0.1358 | (0.1308, 0.1400) |  |
| SD of county random slopes | $\sigma_{v}$ | 0.1011 | (0.0987, 0.1032) |  |
| SD of space-time interaction effects | $\sigma_{\delta}$ | 0.0317 | (0.0302, 0.0329) |  |
| Beta scale | $\phi$ | 533.233 | (516.221,547.795) |  |

Supplementary Table 4. Vaccination statistics (mean, SD, min, max), weeks for which data are available, and number of counties that reported data by US state. For the purposes of our analysis The District of Columbia is a state comprised of 1 county, thus the SD of its vaccination rate by week doesn’t exist (NA). Vaccination data cover February 21, 2021 (week 8) through May 16, 2021 (week 20).

| **State** | **Week Number** | **Total Counties** | **Avg Percent with 1+ dose** | **SD of Percent with 1+ dose** | **Min of Percent with 1+ dose** | **Max of Percent with 1+ dose** |
| --- | --- | --- | --- | --- | --- | --- |
| AK | 14 | 28 | 45.863 | 16.119 | 22.085 | 75.769 |
| AK | 15 | 28 | 47.343 | 14.560 | 22.791 | 73.951 |
| AK | 16 | 28 | 47.020 | 13.637 | 22.631 | 68.528 |
| AK | 17 | 27 | 47.459 | 13.318 | 23.433 | 68.677 |
| AK | 18 | 27 | 47.941 | 13.337 | 23.364 | 68.964 |
| AK | 19 | 27 | 49.049 | 14.321 | 23.346 | 80.461 |
| AK | 20 | 27 | 49.643 | 14.008 | 23.891 | 81.145 |
| AL | 12 | 67 | 17.867 | 3.215 | 8.483 | 24.477 |
| AL | 13 | 67 | 19.670 | 3.408 | 9.707 | 25.971 |
| AL | 16 | 67 | 27.059 | 4.594 | 14.679 | 36.759 |
| AL | 17 | 67 | 27.971 | 4.729 | 15.328 | 38.680 |
| AL | 18 | 67 | 28.729 | 4.793 | 15.881 | 39.646 |
| AL | 19 | 67 | 29.445 | 4.863 | 16.423 | 41.055 |
| AL | 20 | 67 | 29.643 | 4.872 | 16.582 | 41.322 |
| AR | 14 | 75 | 26.030 | 5.604 | 6.654 | 37.873 |
| AR | 15 | 75 | 28.304 | 6.222 | 7.256 | 41.776 |
| AR | 16 | 75 | 29.892 | 6.395 | 8.435 | 43.651 |
| AR | 17 | 75 | 30.399 | 6.352 | 8.425 | 43.359 |
| AR | 18 | 75 | 30.812 | 6.369 | 8.522 | 42.686 |
| AR | 19 | 75 | 31.239 | 6.338 | 9.136 | 43.113 |
| AR | 20 | 75 | 31.453 | 6.183 | 9.146 | 43.313 |
| AZ | 11 | 14 | 23.068 | 8.072 | 9.696 | 39.135 |
| AZ | 12 | 14 | 26.143 | 8.758 | 10.491 | 40.409 |
| AZ | 13 | 14 | 28.669 | 9.332 | 10.802 | 42.935 |
| AZ | 14 | 14 | 31.273 | 10.130 | 11.361 | 44.462 |
| AZ | 15 | 14 | 33.387 | 10.665 | 12.107 | 49.409 |
| AZ | 16 | 14 | 34.857 | 11.028 | 12.461 | 52.854 |
| AZ | 17 | 14 | 36.022 | 11.570 | 12.650 | 56.996 |
| AZ | 18 | 14 | 37.173 | 11.977 | 12.823 | 58.964 |
| AZ | 19 | 14 | 37.977 | 12.216 | 13.004 | 60.616 |
| AZ | 20 | 14 | 38.415 | 12.413 | 13.621 | 62.663 |
| CA | 8 | 58 | 15.994 | 5.044 | 6.946 | 29.853 |
| CA | 9 | 58 | 18.911 | 5.636 | 9.260 | 32.595 |
| CA | 10 | 58 | 22.155 | 6.680 | 11.356 | 46.324 |
| CA | 11 | 58 | 25.736 | 7.354 | 14.242 | 54.473 |
| CA | 12 | 58 | 28.988 | 7.660 | 15.789 | 59.167 |
| CA | 13 | 58 | 32.139 | 8.016 | 17.973 | 61.825 |
| CA | 14 | 58 | 35.472 | 8.860 | 18.955 | 68.291 |
| CA | 15 | 58 | 38.438 | 9.541 | 19.543 | 69.531 |
| CA | 16 | 58 | 40.887 | 10.454 | 20.014 | 71.125 |
| CA | 17 | 58 | 42.796 | 10.995 | 20.584 | 71.922 |
| CA | 18 | 58 | 44.085 | 11.295 | 21.038 | 72.542 |
| CA | 19 | 58 | 45.070 | 11.510 | 21.454 | 72.719 |
| CA | 20 | 57 | 44.756 | 11.039 | 21.467 | 70.113 |
| CO | 14 | 64 | 18.884 | 7.522 | 5.583 | 41.612 |
| CO | 15 | 64 | 23.182 | 8.985 | 6.614 | 52.871 |
| CO | 16 | 64 | 25.713 | 9.365 | 8.666 | 55.155 |
| CO | 17 | 64 | 27.411 | 9.984 | 8.658 | 56.804 |
| CO | 18 | 64 | 28.477 | 10.344 | 9.096 | 57.879 |
| CO | 19 | 64 | 28.896 | 10.597 | 9.379 | 56.857 |
| CO | 20 | 64 | 28.839 | 10.493 | 9.287 | 54.279 |
| CT | 8 | 8 | 15.286 | 2.193 | 13.102 | 19.743 |
| CT | 9 | 8 | 17.661 | 2.354 | 15.084 | 22.365 |
| CT | 10 | 8 | 22.024 | 2.783 | 19.018 | 27.820 |
| CT | 11 | 8 | 25.918 | 2.979 | 22.534 | 32.074 |
| CT | 12 | 8 | 29.583 | 3.149 | 25.993 | 36.216 |
| CT | 13 | 8 | 33.240 | 3.456 | 28.258 | 40.492 |
| CT | 14 | 8 | 40.342 | 3.786 | 34.598 | 48.366 |
| CT | 15 | 8 | 47.912 | 4.648 | 39.766 | 56.921 |
| CT | 16 | 8 | 46.791 | 4.805 | 37.459 | 55.241 |
| CT | 17 | 8 | 48.531 | 5.207 | 38.583 | 57.758 |
| CT | 18 | 8 | 50.471 | 5.356 | 40.439 | 60.096 |
| CT | 19 | 8 | 52.205 | 5.488 | 41.834 | 61.911 |
| DC | 8 | 1 | 10.602 | NA | 10.602 | 10.602 |
| DC | 9 | 1 | 12.779 | NA | 12.779 | 12.779 |
| DC | 10 | 1 | 15.402 | NA | 15.402 | 15.402 |
| DC | 11 | 1 | 19.300 | NA | 19.300 | 19.300 |
| DC | 12 | 1 | 22.580 | NA | 22.580 | 22.580 |
| DC | 13 | 1 | 28.058 | NA | 28.058 | 28.058 |
| DC | 14 | 1 | 32.691 | NA | 32.691 | 32.691 |
| DC | 15 | 1 | 38.264 | NA | 38.264 | 38.264 |
| DC | 16 | 1 | 43.304 | NA | 43.304 | 43.304 |
| DC | 17 | 1 | 47.355 | NA | 47.355 | 47.355 |
| DC | 18 | 1 | 49.691 | NA | 49.691 | 49.691 |
| DC | 19 | 1 | 52.597 | NA | 52.597 | 52.597 |
| DC | 20 | 1 | 53.247 | NA | 53.247 | 53.247 |
| DE | 14 | 3 | 35.618 | 9.934 | 28.123 | 46.885 |
| DE | 15 | 3 | 39.072 | 10.230 | 30.777 | 50.503 |
| DE | 16 | 3 | 42.088 | 10.132 | 33.237 | 53.140 |
| DE | 17 | 3 | 44.398 | 9.394 | 35.552 | 54.258 |
| DE | 18 | 3 | 45.711 | 8.928 | 36.821 | 54.677 |
| DE | 19 | 3 | 47.231 | 8.788 | 38.057 | 55.575 |
| DE | 20 | 3 | 47.541 | 8.691 | 38.364 | 55.647 |
| FL | 8 | 67 | 13.529 | 4.102 | 6.732 | 27.657 |
| FL | 9 | 67 | 15.408 | 4.890 | 7.870 | 30.584 |
| FL | 10 | 67 | 17.499 | 5.245 | 8.919 | 35.026 |
| FL | 11 | 67 | 19.894 | 6.092 | 9.904 | 39.093 |
| FL | 12 | 67 | 22.737 | 7.656 | 11.354 | 54.969 |
| FL | 13 | 67 | 25.058 | 8.201 | 12.594 | 56.778 |
| FL | 14 | 67 | 27.778 | 8.783 | 13.848 | 58.499 |
| FL | 15 | 67 | 30.597 | 9.524 | 14.622 | 60.529 |
| FL | 16 | 67 | 32.704 | 9.593 | 16.915 | 62.029 |
| FL | 17 | 67 | 34.395 | 9.660 | 20.027 | 62.811 |
| FL | 18 | 67 | 35.446 | 9.928 | 20.592 | 63.341 |
| FL | 19 | 67 | 36.423 | 10.185 | 21.024 | 63.931 |
| FL | 20 | 67 | 36.612 | 10.218 | 21.089 | 64.062 |
| GA | 8 | 159 | 11.512 | 3.407 | 2.723 | 22.700 |
| GA | 9 | 159 | 12.599 | 3.592 | 2.961 | 24.832 |
| GA | 10 | 159 | 14.258 | 3.889 | 3.401 | 27.499 |
| GA | 11 | 159 | 16.472 | 4.224 | 4.043 | 30.331 |
| GA | 12 | 159 | 18.671 | 4.573 | 4.960 | 33.272 |
| GA | 13 | 159 | 20.873 | 4.933 | 6.069 | 35.233 |
| GA | 14 | 159 | 22.551 | 5.185 | 7.445 | 37.152 |
| GA | 15 | 159 | 23.818 | 5.399 | 8.233 | 39.170 |
| GA | 16 | 159 | 24.638 | 5.543 | 8.646 | 40.550 |
| GA | 17 | 159 | 25.281 | 5.598 | 9.049 | 41.486 |
| GA | 18 | 159 | 25.839 | 5.680 | 9.398 | 42.373 |
| GA | 19 | 159 | 26.279 | 5.729 | 10.594 | 42.974 |
| GA | 20 | 152 | 26.523 | 5.611 | 10.599 | 43.024 |
| HI | 8 | 4 | 14.937 | 3.298 | 11.214 | 18.940 |
| HI | 9 | 4 | 17.037 | 2.976 | 14.272 | 21.106 |
| HI | 10 | 4 | 20.215 | 3.361 | 17.864 | 25.088 |
| HI | 11 | 4 | 24.537 | 4.292 | 21.596 | 30.912 |
| HI | 12 | 4 | 28.507 | 4.968 | 25.878 | 35.956 |
| HI | 13 | 4 | 31.344 | 4.458 | 28.474 | 37.940 |
| HI | 14 | 4 | 34.451 | 4.066 | 30.868 | 40.296 |
| HI | 15 | 4 | 37.243 | 4.241 | 33.134 | 43.176 |
| HI | 16 | 4 | 39.454 | 4.387 | 35.261 | 45.559 |
| HI | 17 | 4 | 41.058 | 4.293 | 36.952 | 47.046 |
| HI | 18 | 4 | 42.091 | 4.468 | 37.933 | 48.377 |
| HI | 19 | 4 | 42.778 | 4.399 | 38.708 | 48.985 |
| IA | 13 | 99 | 29.569 | 8.062 | 11.605 | 63.345 |
| IA | 14 | 99 | 32.677 | 8.501 | 12.600 | 65.986 |
| IA | 15 | 99 | 35.326 | 9.096 | 14.552 | 75.031 |
| IA | 16 | 99 | 37.630 | 9.416 | 15.333 | 77.672 |
| IA | 17 | 99 | 39.082 | 9.525 | 15.821 | 78.171 |
| IA | 18 | 99 | 39.668 | 9.663 | 15.821 | 78.397 |
| IA | 19 | 99 | 40.352 | 9.784 | 16.192 | 79.082 |
| IA | 20 | 99 | 40.595 | 9.846 | 16.192 | 79.378 |
| ID | 13 | 44 | 22.347 | 5.677 | 14.357 | 44.168 |
| ID | 15 | 44 | 25.709 | 6.391 | 16.180 | 50.628 |
| ID | 16 | 44 | 28.317 | 7.080 | 18.599 | 57.908 |
| ID | 17 | 44 | 29.224 | 7.246 | 19.580 | 60.354 |
| ID | 18 | 44 | 29.923 | 7.349 | 20.198 | 61.535 |
| ID | 19 | 44 | 30.565 | 7.482 | 20.697 | 63.199 |
| ID | 20 | 44 | 30.689 | 7.541 | 20.722 | 63.820 |
| IL | 8 | 102 | 15.114 | 4.000 | 4.270 | 26.550 |
| IL | 9 | 102 | 17.109 | 4.017 | 4.912 | 27.878 |
| IL | 10 | 102 | 19.382 | 3.843 | 6.284 | 28.129 |
| IL | 11 | 102 | 21.839 | 3.841 | 8.262 | 30.079 |
| IL | 12 | 102 | 24.675 | 4.081 | 10.120 | 33.273 |
| IL | 13 | 102 | 26.143 | 4.570 | 11.144 | 35.715 |
| IL | 14 | 102 | 27.513 | 5.058 | 12.047 | 37.591 |
| IL | 15 | 102 | 28.903 | 5.659 | 12.637 | 41.724 |
| IL | 16 | 102 | 30.184 | 6.244 | 13.123 | 45.774 |
| IL | 17 | 102 | 31.084 | 6.679 | 13.435 | 48.669 |
| IL | 18 | 102 | 31.666 | 6.904 | 13.713 | 50.092 |
| IL | 19 | 102 | 32.208 | 7.149 | 13.869 | 51.608 |
| IL | 20 | 102 | 32.316 | 7.203 | 13.886 | 51.872 |
| IN | 8 | 92 | 14.832 | 2.876 | 8.790 | 24.136 |
| IN | 9 | 92 | 16.609 | 3.263 | 9.560 | 26.570 |
| IN | 10 | 92 | 18.782 | 3.815 | 10.852 | 31.183 |
| IN | 11 | 92 | 21.362 | 4.326 | 12.263 | 36.119 |
| IN | 12 | 92 | 23.860 | 4.677 | 13.470 | 41.157 |
| IN | 13 | 92 | 26.358 | 5.011 | 14.555 | 45.174 |
| IN | 14 | 92 | 28.972 | 5.563 | 15.449 | 49.855 |
| IN | 15 | 92 | 31.374 | 6.093 | 16.105 | 52.851 |
| IN | 16 | 92 | 32.712 | 6.406 | 16.583 | 53.719 |
| IN | 17 | 92 | 33.730 | 6.724 | 17.092 | 54.791 |
| IN | 18 | 92 | 34.499 | 6.924 | 17.416 | 55.268 |
| IN | 19 | 92 | 35.137 | 7.090 | 17.665 | 55.830 |
| IN | 20 | 92 | 35.231 | 7.118 | 17.688 | 55.898 |
| KS | 14 | 105 | 37.743 | 8.005 | 19.456 | 55.383 |
| KS | 15 | 105 | 39.755 | 8.195 | 20.262 | 59.314 |
| KS | 16 | 105 | 39.696 | 7.836 | 20.123 | 65.931 |
| KS | 17 | 105 | 39.245 | 7.336 | 19.293 | 61.714 |
| KS | 18 | 105 | 38.884 | 7.632 | 19.027 | 75.363 |
| KS | 19 | 105 | 38.209 | 7.242 | 18.689 | 73.252 |
| KS | 20 | 105 | 38.025 | 7.023 | 18.673 | 71.234 |
| KY | 14 | 120 | 30.091 | 6.432 | 15.186 | 50.581 |
| KY | 15 | 120 | 32.570 | 6.707 | 16.262 | 54.420 |
| KY | 16 | 120 | 33.668 | 6.808 | 16.545 | 56.665 |
| KY | 17 | 120 | 34.215 | 6.865 | 16.854 | 57.305 |
| KY | 18 | 120 | 34.686 | 6.863 | 17.144 | 58.231 |
| KY | 19 | 120 | 35.150 | 7.002 | 17.518 | 58.983 |
| KY | 20 | 120 | 35.336 | 7.020 | 17.780 | 59.073 |
| LA | 11 | 64 | 18.392 | 4.485 | 7.285 | 35.348 |
| LA | 12 | 64 | 20.558 | 5.274 | 7.687 | 42.517 |
| LA | 13 | 64 | 22.620 | 5.879 | 8.547 | 46.364 |
| LA | 14 | 64 | 24.112 | 6.141 | 9.035 | 48.214 |
| LA | 15 | 64 | 25.481 | 6.411 | 9.809 | 50.392 |
| LA | 16 | 64 | 26.286 | 6.587 | 10.326 | 52.634 |
| LA | 17 | 64 | 26.881 | 6.852 | 10.483 | 55.903 |
| LA | 18 | 64 | 27.677 | 7.263 | 10.971 | 60.868 |
| LA | 19 | 64 | 28.693 | 7.593 | 11.243 | 64.979 |
| LA | 20 | 64 | 28.250 | 7.571 | 11.243 | 64.639 |
| MA | 14 | 14 | 28.842 | 14.598 | 0.693 | 40.885 |
| MA | 15 | 14 | 33.093 | 16.881 | 0.830 | 47.081 |
| MA | 16 | 14 | 36.179 | 18.539 | 0.847 | 51.447 |
| MA | 17 | 14 | 39.029 | 20.079 | 0.889 | 56.606 |
| MA | 18 | 14 | 41.058 | 21.223 | 0.929 | 59.537 |
| MA | 19 | 14 | 42.651 | 22.202 | 0.961 | 62.758 |
| MA | 20 | 14 | 42.993 | 22.418 | 1.014 | 63.508 |
| MD | 8 | 24 | 13.907 | 3.228 | 6.980 | 22.160 |
| MD | 9 | 24 | 15.984 | 3.342 | 8.865 | 23.386 |
| MD | 10 | 24 | 18.565 | 3.538 | 11.112 | 25.752 |
| MD | 11 | 24 | 21.444 | 3.798 | 14.118 | 29.128 |
| MD | 12 | 24 | 25.537 | 4.603 | 18.356 | 35.981 |
| MD | 13 | 24 | 29.113 | 5.110 | 21.245 | 40.558 |
| MD | 14 | 24 | 32.596 | 5.562 | 23.072 | 44.450 |
| MD | 15 | 24 | 36.345 | 6.092 | 24.582 | 48.315 |
| MD | 16 | 24 | 39.118 | 6.607 | 26.015 | 51.312 |
| MD | 17 | 24 | 41.678 | 6.983 | 29.173 | 55.249 |
| MD | 18 | 24 | 43.345 | 7.172 | 31.351 | 57.439 |
| MD | 19 | 24 | 44.750 | 7.342 | 33.210 | 59.847 |
| MD | 20 | 24 | 45.573 | 7.567 | 33.842 | 61.374 |
| ME | 8 | 16 | 14.925 | 2.635 | 10.380 | 19.334 |
| ME | 9 | 16 | 17.603 | 2.949 | 12.424 | 22.671 |
| ME | 10 | 16 | 21.128 | 3.282 | 15.197 | 27.028 |
| ME | 11 | 16 | 24.450 | 3.723 | 17.812 | 30.993 |
| ME | 12 | 16 | 27.892 | 3.915 | 21.225 | 35.130 |
| ME | 13 | 16 | 31.332 | 4.155 | 23.911 | 39.326 |
| ME | 14 | 16 | 32.528 | 4.436 | 24.493 | 40.856 |
| ME | 15 | 16 | 42.669 | 4.815 | 35.855 | 50.651 |
| ME | 16 | 16 | 45.708 | 5.527 | 37.677 | 55.900 |
| ME | 17 | 16 | 48.144 | 6.156 | 39.581 | 60.741 |
| ME | 18 | 16 | 49.883 | 6.485 | 40.791 | 63.454 |
| ME | 19 | 16 | 51.182 | 6.553 | 42.132 | 65.002 |
| ME | 20 | 16 | 51.916 | 6.716 | 42.663 | 66.126 |
| MI | 8 | 83 | 16.317 | 4.650 | 9.815 | 29.382 |
| MI | 9 | 83 | 18.740 | 4.759 | 11.102 | 31.768 |
| MI | 10 | 83 | 21.867 | 5.134 | 13.216 | 36.023 |
| MI | 11 | 83 | 25.322 | 5.514 | 15.928 | 38.652 |
| MI | 12 | 83 | 28.366 | 5.901 | 18.346 | 44.488 |
| MI | 13 | 83 | 31.080 | 5.881 | 19.732 | 47.645 |
| MI | 14 | 83 | 33.788 | 6.027 | 21.989 | 51.758 |
| MI | 15 | 83 | 36.345 | 6.156 | 22.884 | 54.965 |
| MI | 16 | 83 | 38.180 | 6.314 | 24.030 | 57.217 |
| MI | 17 | 83 | 39.526 | 6.441 | 24.785 | 59.064 |
| MI | 18 | 83 | 40.417 | 6.506 | 24.965 | 59.924 |
| MI | 19 | 83 | 41.234 | 6.563 | 25.215 | 60.843 |
| MN | 8 | 87 | 16.481 | 5.233 | 7.954 | 39.887 |
| MN | 9 | 87 | 19.712 | 5.371 | 9.972 | 41.625 |
| MN | 10 | 87 | 22.649 | 5.413 | 11.795 | 43.474 |
| MN | 11 | 87 | 22.527 | 5.334 | 11.795 | 43.474 |
| MN | 12 | 87 | 28.580 | 5.818 | 15.667 | 50.778 |
| MN | 13 | 87 | 31.882 | 6.002 | 17.824 | 54.091 |
| MN | 14 | 87 | 34.740 | 6.403 | 20.003 | 60.644 |
| MN | 15 | 87 | 37.543 | 6.502 | 22.390 | 62.786 |
| MN | 16 | 87 | 39.407 | 6.616 | 24.026 | 64.909 |
| MN | 17 | 87 | 40.857 | 6.727 | 25.508 | 65.806 |
| MN | 18 | 87 | 41.876 | 6.833 | 26.442 | 67.051 |
| MN | 19 | 87 | 42.675 | 6.897 | 27.164 | 67.143 |
| MN | 20 | 87 | 42.672 | 6.896 | 27.164 | 67.143 |
| MO | 8 | 114 | 11.488 | 3.475 | 4.490 | 21.248 |
| MO | 9 | 114 | 12.586 | 3.581 | 5.176 | 22.127 |
| MO | 10 | 114 | 14.914 | 4.065 | 5.680 | 24.233 |
| MO | 11 | 114 | 16.853 | 4.432 | 6.942 | 25.685 |
| MO | 12 | 114 | 18.540 | 4.790 | 7.322 | 30.294 |
| MO | 13 | 114 | 19.935 | 5.061 | 8.035 | 31.441 |
| MO | 14 | 115 | 21.409 | 5.398 | 8.807 | 36.559 |
| MO | 15 | 115 | 22.991 | 5.797 | 9.529 | 39.250 |
| MO | 16 | 115 | 23.975 | 6.110 | 10.042 | 42.927 |
| MO | 17 | 115 | 24.677 | 6.396 | 10.516 | 43.644 |
| MO | 18 | 115 | 25.235 | 6.510 | 10.845 | 44.686 |
| MO | 19 | 115 | 25.726 | 6.623 | 11.166 | 45.361 |
| MO | 20 | 115 | 25.917 | 6.673 | 11.320 | 45.831 |
| MS | 11 | 82 | 19.540 | 4.748 | 11.454 | 44.635 |
| MS | 12 | 82 | 21.988 | 5.341 | 12.277 | 49.700 |
| MS | 13 | 82 | 24.245 | 5.718 | 14.846 | 54.320 |
| MS | 14 | 82 | 25.209 | 5.908 | 15.072 | 55.694 |
| MS | 15 | 82 | 27.345 | 6.209 | 16.127 | 59.599 |
| MS | 16 | 82 | 27.832 | 5.373 | 16.127 | 42.157 |
| MS | 17 | 82 | 28.242 | 5.475 | 16.127 | 42.602 |
| MS | 18 | 82 | 29.083 | 5.517 | 19.795 | 43.390 |
| MS | 19 | 82 | 29.729 | 5.513 | 20.098 | 43.973 |
| MS | 20 | 82 | 29.824 | 5.534 | 20.259 | 44.081 |
| MT | 8 | 56 | 15.041 | 3.795 | 7.573 | 24.697 |
| MT | 9 | 56 | 16.828 | 3.832 | 10.216 | 25.969 |
| MT | 10 | 56 | 19.241 | 4.560 | 11.058 | 32.328 |
| MT | 11 | 56 | 21.861 | 4.840 | 12.981 | 32.759 |
| MT | 12 | 56 | 23.899 | 5.220 | 13.822 | 35.777 |
| MT | 13 | 56 | 25.825 | 5.621 | 15.385 | 39.650 |
| MT | 14 | 56 | 28.482 | 6.227 | 16.166 | 43.796 |
| MT | 15 | 56 | 30.415 | 6.915 | 16.526 | 47.396 |
| MT | 16 | 56 | 31.722 | 7.400 | 17.368 | 48.414 |
| MT | 17 | 56 | 32.550 | 7.702 | 17.548 | 48.851 |
| MT | 18 | 56 | 33.184 | 7.844 | 18.570 | 49.912 |
| MT | 19 | 56 | 33.755 | 7.982 | 18.990 | 50.777 |
| NC | 10 | 100 | 18.364 | 3.994 | 9.706 | 31.477 |
| NC | 11 | 100 | 20.529 | 4.221 | 10.688 | 33.927 |
| NC | 12 | 100 | 22.264 | 4.539 | 11.418 | 35.872 |
| NC | 13 | 100 | 24.690 | 4.899 | 12.685 | 39.234 |
| NC | 14 | 100 | 26.015 | 5.198 | 13.305 | 44.237 |
| NC | 15 | 100 | 26.944 | 5.290 | 13.634 | 45.655 |
| NC | 16 | 100 | 29.487 | 5.989 | 14.573 | 48.791 |
| NC | 17 | 100 | 30.034 | 6.020 | 15.042 | 49.342 |
| NC | 18 | 100 | 30.516 | 6.076 | 15.213 | 49.545 |
| NC | 19 | 100 | 30.896 | 6.114 | 15.439 | 49.915 |
| NC | 20 | 100 | 30.970 | 6.126 | 15.479 | 49.947 |
| ND | 15 | 53 | 43.409 | 9.625 | 25.066 | 61.268 |
| ND | 16 | 53 | 44.149 | 9.725 | 25.198 | 63.773 |
| ND | 17 | 53 | 45.076 | 9.666 | 26.385 | 65.230 |
| ND | 18 | 53 | 45.252 | 9.662 | 26.517 | 65.435 |
| ND | 19 | 53 | 45.227 | 9.477 | 27.265 | 63.424 |
| ND | 20 | 53 | 41.631 | 9.036 | 24.941 | 58.180 |
| NE | 14 | 93 | 25.460 | 7.978 | 6.122 | 49.016 |
| NE | 15 | 93 | 28.946 | 8.361 | 7.580 | 53.973 |
| NE | 16 | 93 | 30.691 | 8.237 | 10.062 | 53.974 |
| NE | 17 | 93 | 31.652 | 8.470 | 11.159 | 54.257 |
| NE | 18 | 93 | 31.755 | 8.469 | 10.770 | 52.875 |
| NE | 19 | 93 | 31.469 | 8.548 | 10.244 | 50.307 |
| NE | 20 | 93 | 30.913 | 8.526 | 10.133 | 50.493 |
| NH | 14 | 10 | 41.377 | 9.374 | 27.461 | 56.005 |
| NH | 15 | 10 | 50.823 | 9.446 | 36.075 | 64.847 |
| NH | 16 | 10 | 52.892 | 9.468 | 37.666 | 65.513 |
| NH | 17 | 10 | 53.534 | 9.549 | 38.586 | 66.687 |
| NH | 18 | 10 | 54.628 | 9.828 | 39.619 | 69.098 |
| NH | 19 | 10 | 59.308 | 7.463 | 48.378 | 70.093 |
| NH | 20 | 10 | 59.577 | 7.511 | 48.715 | 70.613 |
| NJ | 8 | 21 | 13.273 | 2.616 | 8.827 | 19.874 |
| NJ | 9 | 21 | 16.781 | 3.060 | 10.819 | 24.773 |
| NJ | 10 | 21 | 20.784 | 3.563 | 13.746 | 29.679 |
| NJ | 11 | 21 | 24.553 | 4.014 | 16.837 | 34.044 |
| NJ | 12 | 21 | 27.959 | 4.349 | 19.849 | 37.373 |
| NJ | 13 | 21 | 31.849 | 4.603 | 23.709 | 41.144 |
| NJ | 14 | 21 | 35.408 | 4.464 | 29.712 | 44.798 |
| NJ | 15 | 21 | 40.343 | 4.446 | 33.449 | 49.892 |
| NJ | 16 | 21 | 43.051 | 4.683 | 35.954 | 53.179 |
| NJ | 17 | 21 | 45.870 | 4.887 | 37.922 | 55.938 |
| NJ | 18 | 21 | 47.598 | 5.038 | 38.996 | 57.647 |
| NJ | 19 | 21 | 49.135 | 5.127 | 40.290 | 59.039 |
| NJ | 20 | 21 | 49.752 | 5.191 | 40.712 | 59.791 |
| NM | 8 | 33 | 19.767 | 6.875 | 7.839 | 35.605 |
| NM | 9 | 33 | 21.919 | 6.555 | 10.717 | 36.605 |
| NM | 10 | 33 | 25.323 | 6.773 | 15.672 | 40.372 |
| NM | 11 | 33 | 26.509 | 6.740 | 16.099 | 40.581 |
| NM | 12 | 33 | 29.179 | 6.781 | 17.872 | 43.163 |
| NM | 13 | 33 | 32.036 | 7.324 | 19.294 | 44.163 |
| NM | 14 | 33 | 35.200 | 8.229 | 20.954 | 51.340 |
| NM | 15 | 33 | 37.611 | 8.876 | 21.733 | 56.188 |
| NM | 16 | 33 | 38.886 | 9.394 | 22.104 | 58.000 |
| NM | 17 | 33 | 43.432 | 11.249 | 23.708 | 63.452 |
| NM | 18 | 33 | 44.321 | 11.307 | 24.232 | 64.056 |
| NM | 19 | 33 | 45.257 | 11.506 | 24.741 | 65.925 |
| NV | 8 | 16 | 12.093 | 6.850 | 4.156 | 27.525 |
| NV | 9 | 16 | 13.236 | 7.139 | 4.864 | 28.502 |
| NV | 10 | 16 | 14.547 | 7.295 | 5.000 | 29.279 |
| NV | 11 | 16 | 16.176 | 7.860 | 5.000 | 30.211 |
| NV | 12 | 16 | 17.972 | 8.648 | 5.000 | 31.321 |
| NV | 13 | 17 | 23.384 | 5.984 | 9.095 | 31.942 |
| NV | 14 | 17 | 24.600 | 6.494 | 9.896 | 33.756 |
| NV | 15 | 17 | 26.839 | 7.356 | 11.181 | 38.411 |
| NV | 16 | 17 | 28.575 | 7.491 | 12.491 | 40.981 |
| NV | 17 | 17 | 29.914 | 8.033 | 13.437 | 43.476 |
| NV | 18 | 17 | 30.817 | 8.246 | 14.189 | 44.528 |
| NV | 19 | 17 | 31.589 | 8.484 | 14.407 | 45.699 |
| NV | 20 | 17 | 31.783 | 8.535 | 14.528 | 46.083 |
| NY | 8 | 62 | 13.925 | 4.508 | 7.685 | 39.380 |
| NY | 9 | 62 | 17.304 | 4.941 | 10.534 | 44.316 |
| NY | 10 | 62 | 21.924 | 5.367 | 14.450 | 49.592 |
| NY | 11 | 62 | 25.408 | 5.551 | 17.078 | 52.785 |
| NY | 12 | 62 | 28.898 | 5.687 | 20.299 | 56.024 |
| NY | 13 | 62 | 32.114 | 5.777 | 22.950 | 57.767 |
| NY | 14 | 62 | 35.539 | 6.108 | 25.808 | 59.964 |
| NY | 15 | 62 | 39.103 | 6.544 | 28.737 | 60.915 |
| NY | 16 | 62 | 40.770 | 6.748 | 29.644 | 61.889 |
| NY | 17 | 62 | 43.950 | 7.142 | 31.154 | 63.179 |
| NY | 18 | 62 | 45.461 | 7.352 | 31.785 | 64.017 |
| NY | 19 | 62 | 46.646 | 7.457 | 32.533 | 64.583 |
| NY | 20 | 62 | 47.137 | 7.522 | 32.655 | 64.629 |
| OH | 8 | 88 | 13.408 | 1.993 | 7.502 | 19.371 |
| OH | 9 | 88 | 16.081 | 2.240 | 7.987 | 22.290 |
| OH | 10 | 88 | 19.371 | 2.564 | 8.744 | 27.154 |
| OH | 11 | 88 | 22.529 | 2.914 | 10.027 | 31.808 |
| OH | 12 | 88 | 25.462 | 3.649 | 10.507 | 36.168 |
| OH | 13 | 88 | 28.486 | 4.500 | 11.504 | 39.985 |
| OH | 14 | 88 | 30.918 | 5.218 | 12.236 | 45.144 |
| OH | 15 | 88 | 32.579 | 5.784 | 12.803 | 49.582 |
| OH | 16 | 88 | 33.576 | 6.171 | 13.007 | 51.738 |
| OH | 17 | 88 | 34.385 | 6.439 | 13.246 | 53.176 |
| OH | 18 | 88 | 35.104 | 6.617 | 13.560 | 54.282 |
| OH | 19 | 88 | 35.666 | 6.758 | 13.813 | 55.178 |
| OH | 20 | 88 | 35.753 | 6.779 | 13.860 | 55.410 |
| OK | 14 | 77 | 30.638 | 5.641 | 19.664 | 45.547 |
| OK | 15 | 77 | 31.557 | 5.669 | 20.969 | 46.040 |
| OK | 16 | 77 | 31.834 | 5.787 | 20.956 | 47.609 |
| OK | 17 | 77 | 31.909 | 5.830 | 21.091 | 47.485 |
| OK | 18 | 77 | 32.088 | 5.814 | 21.666 | 47.716 |
| OK | 19 | 77 | 32.435 | 5.885 | 22.044 | 47.837 |
| OK | 20 | 77 | 32.628 | 5.876 | 21.996 | 47.970 |
| OR | 8 | 36 | 13.251 | 4.085 | 8.572 | 33.333 |
| OR | 9 | 36 | 15.324 | 3.985 | 10.156 | 33.934 |
| OR | 10 | 36 | 18.502 | 3.896 | 12.660 | 35.210 |
| OR | 11 | 36 | 21.436 | 4.504 | 15.548 | 37.237 |
| OR | 12 | 36 | 24.795 | 6.706 | 16.679 | 55.929 |
| OR | 13 | 36 | 26.819 | 8.094 | 17.909 | 66.547 |
| OR | 15 | 36 | 33.117 | 6.191 | 22.906 | 49.089 |
| OR | 16 | 36 | 35.226 | 6.770 | 23.695 | 50.932 |
| OR | 17 | 36 | 37.312 | 7.558 | 24.517 | 54.120 |
| OR | 18 | 36 | 38.771 | 8.045 | 25.270 | 56.503 |
| OR | 19 | 36 | 40.333 | 8.606 | 26.244 | 58.412 |
| OR | 20 | 36 | 40.859 | 8.801 | 26.526 | 59.919 |
| PA | 8 | 66 | 13.166 | 4.831 | 6.045 | 33.752 |
| PA | 9 | 66 | 15.157 | 5.139 | 6.758 | 35.957 |
| PA | 10 | 66 | 17.335 | 5.363 | 7.268 | 37.345 |
| PA | 11 | 66 | 20.494 | 5.588 | 8.775 | 39.259 |
| PA | 12 | 66 | 23.064 | 5.654 | 11.721 | 41.816 |
| PA | 13 | 66 | 26.435 | 5.975 | 14.069 | 44.619 |
| PA | 14 | 66 | 29.012 | 6.314 | 14.892 | 47.581 |
| PA | 15 | 66 | 32.345 | 7.345 | 15.594 | 51.966 |
| PA | 16 | 66 | 34.927 | 7.695 | 16.677 | 52.863 |
| PA | 17 | 67 | 38.143 | 7.575 | 20.427 | 54.561 |
| PA | 18 | 67 | 40.049 | 7.998 | 21.087 | 56.766 |
| PA | 19 | 67 | 41.489 | 8.349 | 21.562 | 60.409 |
| PA | 20 | 67 | 41.971 | 8.492 | 21.741 | 61.733 |
| RI | 14 | 5 | 39.374 | 4.076 | 32.435 | 42.287 |
| RI | 15 | 5 | 44.362 | 4.224 | 37.023 | 47.471 |
| RI | 16 | 5 | 49.261 | 4.801 | 40.928 | 52.815 |
| RI | 17 | 5 | 53.864 | 5.133 | 44.860 | 57.443 |
| RI | 18 | 5 | 55.848 | 5.145 | 46.797 | 59.021 |
| RI | 19 | 5 | 57.491 | 4.921 | 48.860 | 60.845 |
| RI | 20 | 5 | 58.316 | 5.143 | 49.249 | 61.787 |
| SC | 14 | 46 | 28.304 | 7.095 | 16.303 | 54.236 |
| SC | 15 | 46 | 30.532 | 7.254 | 17.901 | 55.558 |
| SC | 16 | 46 | 31.512 | 7.120 | 18.649 | 55.733 |
| SC | 17 | 46 | 32.404 | 7.093 | 19.335 | 55.016 |
| SC | 18 | 46 | 33.054 | 6.966 | 20.106 | 54.334 |
| SC | 19 | 46 | 33.689 | 6.913 | 21.112 | 54.102 |
| SC | 20 | 46 | 33.852 | 6.855 | 21.268 | 53.734 |
| SD | 14 | 65 | 29.182 | 12.524 | 1.346 | 55.887 |
| SD | 15 | 65 | 31.044 | 13.175 | 1.484 | 57.586 |
| SD | 16 | 65 | 31.968 | 13.572 | 1.503 | 60.102 |
| SD | 17 | 65 | 32.628 | 13.827 | 1.582 | 61.119 |
| SD | 18 | 65 | 33.177 | 14.025 | 1.611 | 62.863 |
| SD | 19 | 65 | 33.507 | 14.130 | 1.631 | 63.154 |
| SD | 20 | 65 | 33.587 | 14.145 | 1.651 | 63.234 |
| TN | 8 | 95 | 12.189 | 2.624 | 6.302 | 18.278 |
| TN | 9 | 95 | 14.312 | 2.840 | 7.954 | 21.285 |
| TN | 10 | 95 | 16.685 | 3.227 | 9.835 | 24.838 |
| TN | 11 | 95 | 19.132 | 3.607 | 11.498 | 28.007 |
| TN | 12 | 95 | 21.631 | 4.126 | 12.837 | 32.647 |
| TN | 13 | 95 | 23.983 | 4.663 | 13.714 | 36.130 |
| TN | 14 | 95 | 25.537 | 5.045 | 14.237 | 38.726 |
| TN | 15 | 95 | 27.808 | 5.594 | 15.192 | 42.586 |
| TN | 16 | 95 | 28.902 | 5.893 | 15.607 | 44.356 |
| TN | 17 | 95 | 29.693 | 6.079 | 15.930 | 45.481 |
| TN | 18 | 95 | 30.357 | 6.231 | 16.207 | 46.509 |
| TN | 19 | 95 | 31.059 | 6.398 | 16.377 | 47.486 |
| TN | 20 | 95 | 31.364 | 6.509 | 16.438 | 47.961 |
| TX | 8 | 254 | 12.626 | 5.236 | 3.743 | 35.152 |
| TX | 9 | 254 | 13.499 | 5.270 | 4.153 | 34.545 |
| TX | 10 | 254 | 17.732 | 5.732 | 7.411 | 39.913 |
| TX | 11 | 254 | 20.768 | 6.111 | 8.906 | 54.922 |
| TX | 12 | 254 | 22.821 | 6.850 | 10.216 | 68.737 |
| TX | 13 | 254 | 24.862 | 7.442 | 11.414 | 68.789 |
| TX | 14 | 254 | 26.406 | 7.368 | 12.330 | 78.549 |
| TX | 15 | 254 | 29.605 | 8.122 | 12.735 | 95.000 |
| TX | 16 | 254 | 30.683 | 8.383 | 13.526 | 95.000 |
| TX | 17 | 254 | 31.469 | 7.943 | 13.805 | 76.831 |
| TX | 18 | 254 | 32.265 | 7.431 | 14.377 | 70.615 |
| TX | 19 | 254 | 32.696 | 7.596 | 14.331 | 72.658 |
| TX | 20 | 254 | 32.796 | 7.659 | 13.235 | 72.718 |
| UT | 14 | 29 | 31.289 | 7.091 | 24.335 | 49.454 |
| UT | 15 | 29 | 35.415 | 8.110 | 26.600 | 54.681 |
| UT | 16 | 29 | 36.779 | 8.315 | 27.375 | 57.529 |
| UT | 17 | 29 | 37.947 | 8.301 | 28.330 | 61.561 |
| UT | 18 | 29 | 38.555 | 8.246 | 29.020 | 64.287 |
| UT | 19 | 29 | 39.026 | 8.456 | 28.522 | 66.658 |
| UT | 20 | 29 | 38.546 | 8.222 | 28.335 | 66.705 |
| VA | 10 | 133 | 19.327 | 4.770 | 4.006 | 33.000 |
| VA | 11 | 133 | 22.344 | 5.046 | 4.995 | 37.678 |
| VA | 12 | 133 | 25.365 | 5.431 | 6.046 | 42.436 |
| VA | 13 | 133 | 28.841 | 5.840 | 7.791 | 54.760 |
| VA | 14 | 133 | 31.949 | 6.353 | 9.916 | 66.123 |
| VA | 15 | 133 | 34.852 | 7.075 | 11.703 | 78.298 |
| VA | 16 | 133 | 37.110 | 7.672 | 12.976 | 85.919 |
| VA | 17 | 133 | 38.914 | 8.243 | 14.212 | 93.952 |
| VA | 18 | 133 | 40.305 | 8.367 | 15.570 | 95.000 |
| VA | 19 | 133 | 41.443 | 8.475 | 16.449 | 95.000 |
| VA | 20 | 133 | 41.875 | 8.491 | 16.770 | 95.000 |
| VT | 9 | 14 | 15.107 | 1.634 | 11.569 | 18.302 |
| VT | 10 | 14 | 17.995 | 1.803 | 14.068 | 21.318 |
| VT | 11 | 14 | 22.095 | 2.305 | 17.897 | 26.585 |
| VT | 12 | 14 | 25.839 | 2.823 | 19.877 | 30.635 |
| VT | 13 | 14 | 29.443 | 3.260 | 22.116 | 34.772 |
| VT | 14 | 14 | 33.954 | 3.136 | 28.428 | 39.274 |
| VT | 15 | 14 | 38.534 | 3.691 | 31.186 | 43.771 |
| VT | 16 | 14 | 43.183 | 4.013 | 36.557 | 48.386 |
| VT | 17 | 14 | 47.415 | 4.873 | 38.325 | 54.928 |
| VT | 18 | 14 | 51.342 | 5.694 | 40.662 | 60.426 |
| VT | 19 | 14 | 55.245 | 6.286 | 43.388 | 65.043 |
| WA | 8 | 39 | 13.997 | 4.727 | 7.243 | 26.372 |
| WA | 9 | 39 | 15.751 | 5.100 | 7.893 | 29.769 |
| WA | 10 | 39 | 17.655 | 5.336 | 9.571 | 32.454 |
| WA | 11 | 39 | 17.601 | 5.360 | 9.571 | 32.454 |
| WA | 12 | 39 | 23.523 | 6.308 | 12.972 | 42.829 |
| WA | 13 | 39 | 26.147 | 6.969 | 15.526 | 47.469 |
| WA | 14 | 39 | 28.775 | 7.481 | 17.486 | 50.601 |
| WA | 15 | 39 | 31.559 | 7.945 | 18.778 | 53.648 |
| WA | 16 | 39 | 33.694 | 8.460 | 19.910 | 56.597 |
| WA | 17 | 39 | 35.561 | 8.948 | 20.629 | 59.259 |
| WA | 18 | 39 | 37.071 | 9.794 | 21.366 | 64.828 |
| WA | 19 | 39 | 38.393 | 10.017 | 21.483 | 65.823 |
| WA | 20 | 39 | 38.708 | 10.031 | 21.663 | 65.982 |
| WI | 11 | 72 | 23.838 | 4.288 | 14.177 | 37.335 |
| WI | 12 | 72 | 27.413 | 4.664 | 17.116 | 42.611 |
| WI | 13 | 72 | 30.712 | 5.141 | 18.109 | 45.970 |
| WI | 14 | 72 | 33.993 | 5.686 | 19.869 | 52.064 |
| WI | 15 | 72 | 35.934 | 5.921 | 20.764 | 54.612 |
| WI | 17 | 72 | 40.372 | 6.486 | 23.767 | 59.017 |
| WI | 18 | 72 | 41.346 | 6.580 | 24.662 | 60.677 |
| WI | 19 | 72 | 42.157 | 6.672 | 25.213 | 61.785 |
| WI | 20 | 72 | 42.506 | 6.714 | 25.345 | 62.600 |
| WV | 12 | 55 | 24.911 | 4.131 | 15.813 | 39.699 |
| WV | 13 | 55 | 27.327 | 4.507 | 17.871 | 42.521 |
| WV | 14 | 55 | 28.526 | 4.573 | 19.358 | 42.974 |
| WV | 15 | 55 | 28.526 | 4.573 | 19.358 | 42.974 |
| WV | 16 | 55 | 35.082 | 5.387 | 23.438 | 48.706 |
| WV | 17 | 55 | 35.778 | 5.466 | 23.830 | 48.940 |
| WV | 18 | 55 | 36.422 | 5.517 | 24.236 | 49.379 |
| WV | 19 | 55 | 36.878 | 5.533 | 24.492 | 49.700 |
| WV | 20 | 55 | 38.726 | 5.497 | 24.594 | 52.478 |
| WY | 8 | 23 | 14.863 | 2.880 | 7.926 | 21.075 |
| WY | 9 | 23 | 16.175 | 3.505 | 8.690 | 22.844 |
| WY | 11 | 23 | 20.968 | 4.704 | 12.209 | 34.210 |
| WY | 12 | 23 | 22.276 | 5.437 | 12.742 | 39.290 |
| WY | 14 | 23 | 24.898 | 8.139 | 13.992 | 55.728 |
| WY | 15 | 23 | 26.131 | 8.750 | 14.678 | 58.848 |
| WY | 16 | 23 | 27.011 | 9.568 | 15.155 | 63.476 |
| WY | 17 | 23 | 27.593 | 9.863 | 15.518 | 65.087 |
| WY | 18 | 23 | 27.593 | 9.863 | 15.518 | 65.087 |
| WY | 19 | 23 | 27.593 | 9.863 | 15.518 | 65.087 |
